# Psilocybin mushrooms and public health in Brazil: a low-risk adverse event profile calls for evidence-based regulatory discussions

**DOI:** 10.1101/2024.07.11.24310147

**Authors:** Marcel Nogueira, Solimary García-Hernández, Gleicy Sotéro Roberto, Leonardo Zanella Marques

**Affiliations:** Scientific Department, Instituto Micélio Sagrado, São Paulo, Brazil; Department of Ecology, Biosciences Institute, University of São Paulo, São Paulo, Brazil; Department of Clinical and Toxicological Analyses, School of Pharmaceutical Sciences, University of São Paulo, São Paulo, Brazil; Interdisciplinary Cooperation for Ayahuasca Research and Outreach (ICARO), School of Medical Sciences, University of Campinas, Campinas, São Paulo, Brazil

**Keywords:** Psilocybin, Psilocybe cubensis, Psychedelics, Drug policy, Drug abuse, Adverse events

## Abstract

**Background:** Current drug policy classifies psilocybin, a substance produced by psychoactive mushrooms, as having a high potential for abuse, neglecting its therapeutic properties. We aimed to investigate if psilocybin mushrooms pose a risk to Brazilian public health compared to other toxic agents and whether evidence-based regulatory discussions are needed.

**Methods:** A retrospective cross-sectional study was conducted following STROBE guidelines. Data were obtained from the Sistema de Agravos de Notificação (SINAN) on adverse events reported from 2007 to 2022. Participants were categorized into three groups: drug abuse, psilocybin mushrooms, and unknown mushrooms. Clinical outcomes assessed included non-hospitalization, hospitalization, and death. Associations between variables were analyzed using the Chi-square test.

**Results:** A total of 112,451 individuals sought medical attention for drug abuse-related adverse events. Among them, men constituted the majority (n = 79,514; 70.7%), followed by whites (n = 37,565; 33.4%) and those aged 26-35 (n = 29,163; 25.9%) (p < 0.001). Alcohol was the primary toxic agent (n = 71,824; 49.2%) (p < 0.001). The psilocybin mushroom group reported 13 adverse events, and the unknown mushroom group recorded 51 adverse events. Hospitalization rates were 19.5% (n = 21,923) for drug abuse, 46.2% (n = 6) for psilocybin mushrooms (0.02% of all hospitalizations) (99% CI: 10.6% - 81.6%), and 23.5% (n = 12) for unknown mushrooms (0.04% of hospitalizations) (99% CI: 8.3% - 38.7%). The mortality rate was 1.8% (n = 2035) for drug abuse, with no fatalities in the psilocybin or unknown mushroom groups. Most hospitalizations involved alcohol (45.0%), and deaths were mainly associated with cocaine (33.3%).

**Conclusion:** Our findings suggest that psilocybin mushrooms have a low-risk profile for adverse events, although underreporting may be a factor. This study highlights the need for evidence-based regulatory discussions to prevent arbitrary arrests and ensure safe access to psilocybin for clinical and ceremonial use.

## Introduction

Psilocybin mushrooms and other psychedelics have a long history of use, dating back to prehistoric times in some civilizations (Carod-Artal, 2015; Guerra-Doce, 2015). Even after more than 50 years of prohibition in several countries, the use of psychedelics remained clandestine, often associated with spiritual retreats located in regions with less restrictive laws (Rucker & Young, 2021). This clandestine use spurred renewed scientific interest in their clinical applications in psychiatry, especially psilocybin — the primary serotonergic alkaloid agonist produced by psilocybin mushrooms (Irizarry et al., 2022). Due to its fast-acting antidepressant properties, crucial for regulating mood, anxiety, and perception (Irizarry et al., 2022), consumption has increased in the last decade (Australian Institute of Health and Welfare, 2020).

Supporting these claims, an observational study focusing on the benefits of psilocybin mushrooms gathered data from 1368 people who self-administered these fungi for mental health disorders and specific life concerns. The study revealed positive changes in aspects of well-being, psychiatric symptoms, social-emotional skills, and healthy behaviors (Kopra et al., 2023). Moreover, other studies on synthetic psilocybin demonstrate improved quality of life and prolonged reductions in symptoms of anxiety and depression after a single dose (Goodwin et al., 2022, 2023; Griffiths et al., 2016; Irizarry et al., 2022).

Although it is not yet clear whether the benefits of psilocybin are generalizable to larger populations (Irizarry et al., 2022), public pressure and lobbying have influenced regulatory changes in some countries. In Australia, these efforts resulted in its legalization in 2023 (Haridy, 2023). Conversely, in the United States (USA), drug policy (Controlled Substances Act) classifies psilocybin as dangerous, without therapeutic application, and with a high potential for abuse. Experts point out that this classification was carried out in the absence of any pharmacological, neuroscientific, or psychiatric evidence (Levin et al., 2022). Despite this, the regulation of psilocybin still varies in some parts of the country, such as the state of Oregon, which adopted a progressive policy by allowing the therapeutic use of this substance (Holoyda B, 2023).

Additionally, the hegemonic influence of the USA on international drug policy determines the classification system of regulatory agencies (Levin et al., 2022). In Brazil, for example, regulation of psilocybin mushrooms is marked by ambiguities that hinder both research and its clinical application. The Brazilian Health Regulatory Agency (ANVISA) maintains psilocybin and psilocin (its active metabolite) as prohibited substances but does not mention any species of psilocybin mushrooms on the list of proscribed plants and fungi (BRASIL, 1998). This gap allows companies specializing in psilocybin mushrooms, such as *Psilocybe cubensis*, to operate in a “gray area” since the early 1990s, with publicly available information about their operations. Because of regulatory uncertainties surrounding psilocybin mushrooms and growing public interest, Brazilian media has reported arbitrary arrests of businesspeople in recent years. It is believed that such arbitrariness occurs both because of insufficient information about the nature and applicability of the fungus, and due to a lack of constitutionally adequate understanding on the subject by public security bodies and the Brazilian criminal justice system, highlighting the need for clearer and evidence-based regulation (BBC News Brasil, 2023; UOL, 2023).

Considering the regulatory ambiguity in Brazil, it is important to note that psilocybin mushrooms are naturally occurring fungi, primarily from the genus *Psilocybe*, which comprises approximately 50% of psychoactive species found in subtropical and humid regions, including the Brazilian biome (Plazas & Faraone, 2023). As they are not prohibited or under special control by ANVISA (BRASIL, 1998), there is no official information about adverse events (Appendix A). Given the current drug policy, which overlooks the therapeutic properties of psilocybin (BRASIL, 1998; Levin et al., 2022), we investigate if psilocybin mushrooms pose a risk to Brazilian public health compared to other toxic agents and whether evidence-based regulatory discussions are needed.

Therefore, our objectives were:

1. To analyze the demographic profile of individuals seeking emergency medical attention in Brazil due to adverse events from drug abuse, identifying occurrences involving psilocybin and unknown mushrooms.
2. To estimate the prevalence of non-hospitalization, hospitalization and death associated with drug abuse, including psilocybin and unknown mushrooms.
3. To estimate overall prevalence rates between toxic agents for each of these outcomes.

## Methods Study design

Retrospective cross-sectional study conducted in accordance with the STROBE Initiative (von Elm et al., 2008). We use anonymized and publicly available information. No additional ethical approvals were required.

### Setting

This study examined emergency department visits in Brazil for adverse events related to drug abuse, including hospital admissions and outpatient clinic visits, from 2007 to 2022.

### Participants

We only included participants whose adverse event was reported as “drug abuse.” Participants involved in circumstances such as suicide attempt, accidental ingestion, habitual use, food ingestion, self-medication, therapeutic use, administration error, environmental intoxication, homicide, abortion attempt, medical prescription and not informed were excluded.

### Outcomes

Participant outcomes were assessed, with a focus on the prevalence of non-hospitalization, hospitalization, and death following adverse events associated with drug abuse.

### Data Sources/Measurement

Data were obtained from the Sistema de Informação de Agravos de Notificação (SINAN), a database maintained by the Departamento de Informação e Informática do Sistema Único de Saúde (DATASUS), the Brazilian Unified Health System’s information technology department. The SINAN system plays a crucial role in monitoring Brazilian public health by systematically collecting information on notifiable diseases and adverse events nationwide, including data from emergency medical services. Healthcare professionals from both public and private sectors contribute data to SINAN, which is then transmitted through a computerized network. This integrated system enables timely surveillance, analysis, and response to health issues, supporting effective public health interventions and providing a comprehensive overview of the country’s health status.

Our focus is on records of “exogenous intoxication” in Brazil from 2007 to 2022. This period was chosen because the information was only available for these years at the time of data collection, and there were no data available for the preceding years. Exogenous intoxication refers to adverse events caused by exposure to chemical or toxic agents. These agents can enter the body through ingestion, inhalation, skin absorption, or contact with mucous membranes, leading to a range of symptoms from mild to severe and potentially fatal conditions. Examples include medications, drugs, chemicals, plants, pesticides, and others (Lisboa et al., 2023).

The data collection was conducted in July 2023 and evaluated year by year using the TabWin software. Subsequently, data was transferred to Microsoft Excel for filtering, focusing solely on cases of drug abuse. Due to a significant number of erroneous records and inconsistencies in the nomenclature of reported substances, MN standardized the data as follows:

1. Substance name conversion: ethyl alcohol, alcoholic beverages, street drug names, and plant-based drugs were converted to their scientific nomenclature.
2. Error correction: duplicates, typos, and other inaccuracies in substance names (misspellings, incorrect abbreviations) were corrected.
3. Unknown substance classification: terms like “drugs,” “illegal drug,” “unknown medicine,” “plant,” and “toxic plant” were all categorized as “unknown substance.”
4. Mushroom standardization: “mushroom tea (psilocybin)” and “*Psilocybe cubensis*” were categorized as “psilocybin mushrooms,” while “toxic mushroom” or simply “mushroom” were categorized as “unknown mushroom.”
5. Medication Standardization: medication names were standardized using the Anatomical Therapeutic Chemical (ATC) classification system, and combination drugs were stratified by their individual active ingredients.

Following these steps, the sample was divided into three groups: drug abuse, psilocybin mushrooms, and unknown mushrooms.

### Bias

To ensure the accuracy and consistency of the data analyzed in this study, an initial statistical verification was conducted by the authors. Additionally, to strengthen the reliability of the results, the statistical analysis was reviewed by an independent expert. The review confirmed the accuracy of the data and the appropriateness of the methods used, demonstrating the robustness of the study.

### Sample size

We present a representative sample of emergency department visits due to drug abuse from a historical series. Since there is no official data on psilocybin mushrooms consumption in Brazil, and considering the hypothesis that adverse events from these fungi are rare (Winstock A et al., 2017), we searched for psilocybin-related entries within the existing drug abuse information (DATASUS) to analyze the preliminary estimates of the prevalence in our population.

To ensure that the preliminary estimates of the prevalence accurately reflected a low-risk adverse event profile, we calculated the minimum sample size required for a margin of error of 0.01% and a confidence level of 95%. Following the methodological guidelines proposed by (Chow et al., 2017; Kish L, 1965; Lohr, 2010), we applied the proportion formula:

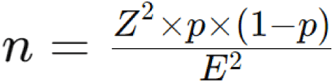

Where:

“n” = required sample size.

“Z” = Z-score for the desired confidence level.

“p” = estimated proportion of psilocybin-related entries within existing drug abuse information.

*E* = margin of error.

### Variables

We used frequency and percentage to describe the demographic characteristics. These variables include the following distributions: sex, age group (<18, 18-25, 26-35, 36-45, 46-55, 56-65, 65+), race, toxic agent, number of toxic agents administered per participant, diagnostic criteria, type of exposure, final classification, and clinical evolution.

The age groups were chosen to represent distinct life stages and potential differences in susceptibility to adverse events. Presenting the data as frequencies and percentages allows for a straightforward comparison of these demographic characteristics across all groups.

### Statistical analysis

Data analysis was performed through RStudio version 4.3 and Microsoft Excel 365. To assess associations between variables, we employed the chi-square test as part of inferential statistics, with a statistical significance level of ≤ 5%. In this retrospective cross-sectional study, we utilized a 99% confidence interval (CI) to compare the prevalence rates of outcomes between psilocybin and unknown mushroom groups due to the small sample sizes. This decision was made to account for the rarity of the outcome and provide a more conservative estimate of variability, given the limitations of the study design.

## Results

### Participants

In the years 2007 to 2022, 924,736 individuals sought medical attention for the adverse event “exogenous intoxication”. Among the circumstances involved, suicide attempts (n = 367,061), accidental intoxication (n = 171,247) and drug abuse (n = 112,451) stood out. All drug abuse cases were included in our study (Figure 1). We then separated participants into three groups: drug abuse (n = 112,451), psilocybin mushrooms (n = 13), and unknown mushrooms (n = 51).

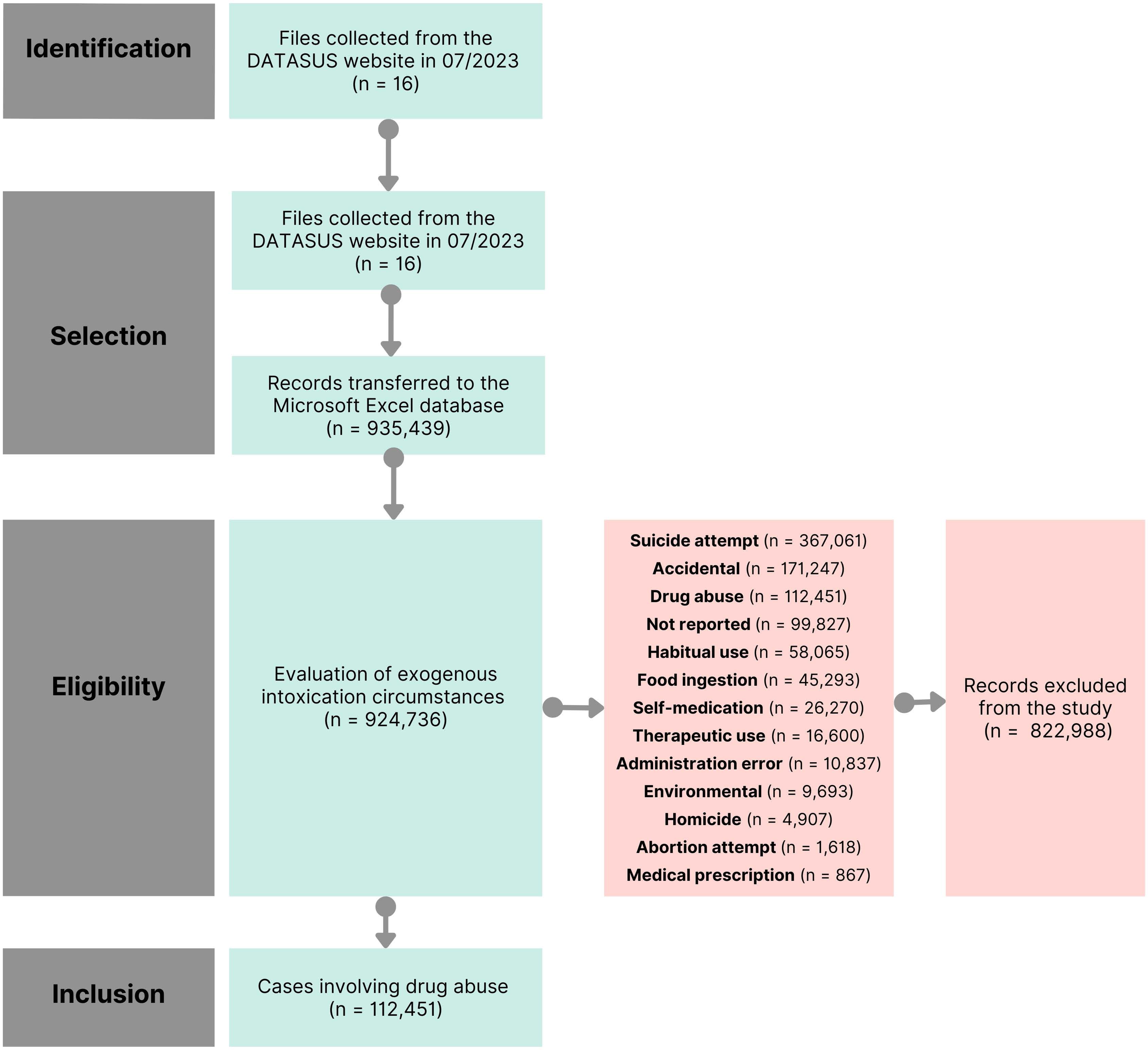

### Descriptive data

#### Demographic characteristics of drug abuse group

In analyzing the 112,451 adverse events due to drug abuse, all categorical variables showed a p-value < 0.001 (Table 1). The most represented demographics were men (n = 79,514; 70.7%), the age group 26-35 (n = 29,163; 25.9%), and white individuals (n = 37,565; 33.4%). We classified 18 toxic agents (Table 2), with alcohol being the most prevalent (n = 71,824; 49.2%) (p < 0.001). Among medications (n = 14,794; 10.1%), antiepileptics were statistically significant (p < 0.001) (Appendix B). In 69.2% (n = 77,768) of cases, a single toxic agent was administered. The most common adverse event confirmation criterion was clinical (n = 65,079; 57.9%), with single-dose acute exposure (n = 42,788; 38.1%) and the final classification “confirmed intoxication” (n = 88,595; 78.8%) being the most frequent. Missing data included race (n = 28,769; 25.6%), age group (n = 3,694; 3.3%), sex (n = 16; 0%), and type of exposure (n = 24,634; 21.9%).

**Table 1.**
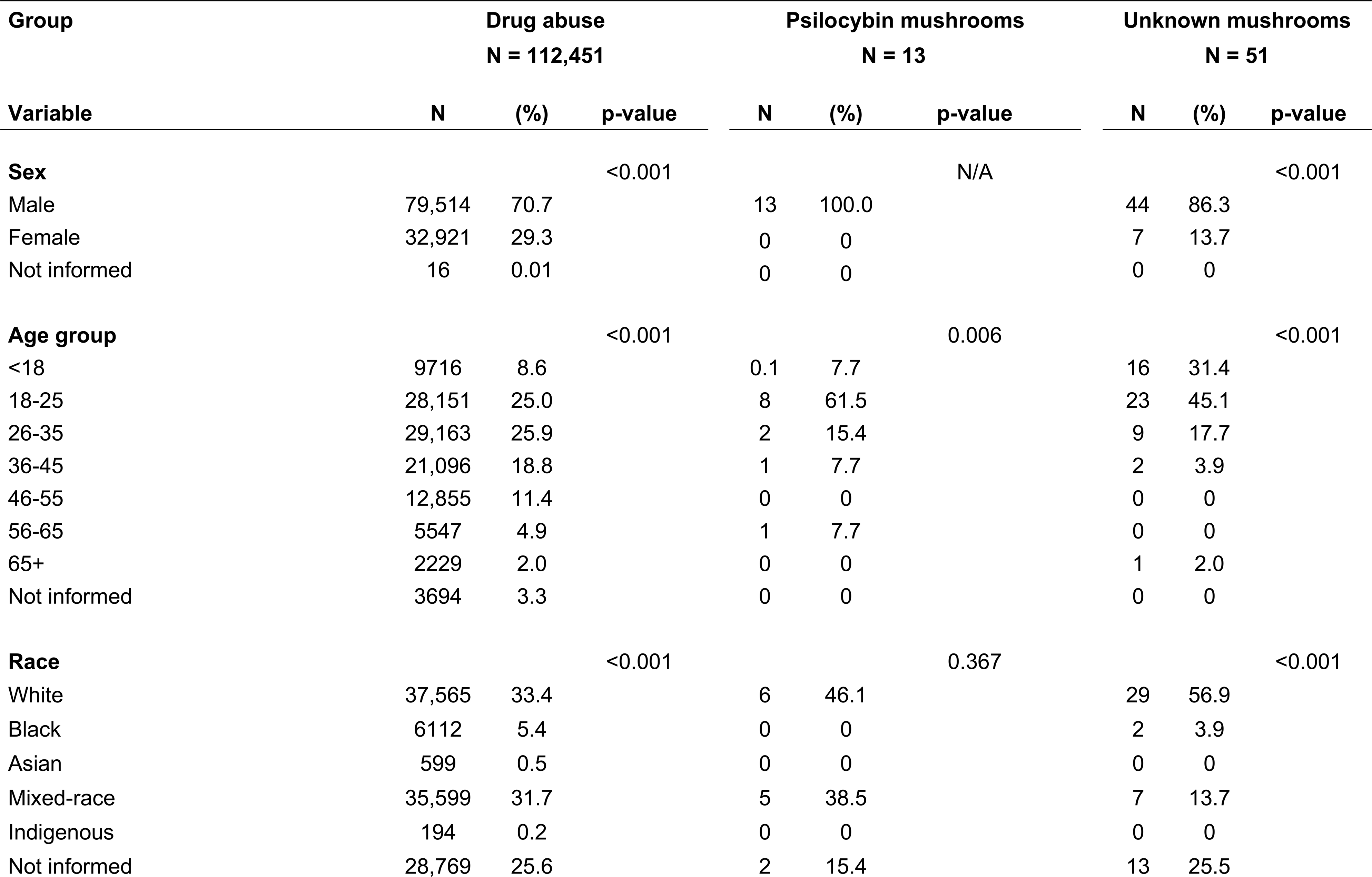
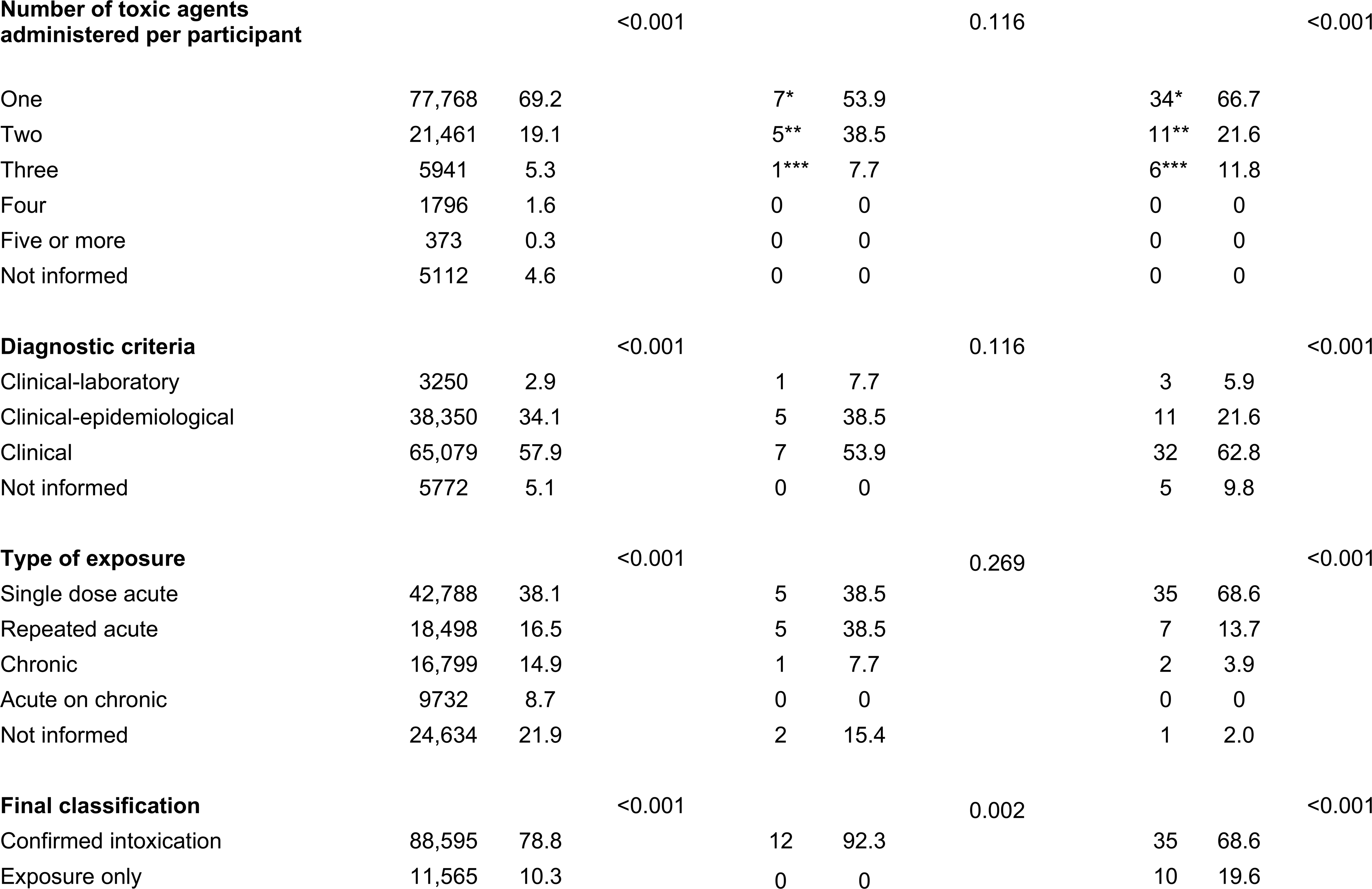
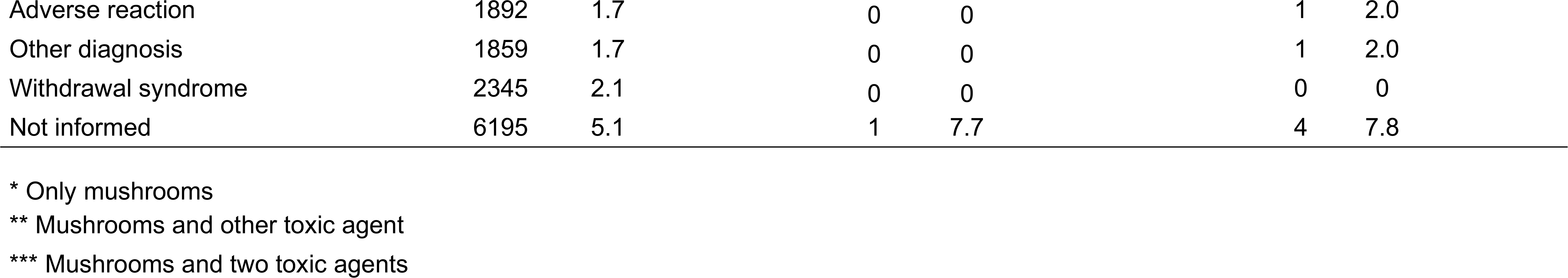
Demographic characteristics of adverse events by group: drug abuse, psilocybin mushrooms and unknown mushrooms.

**Table 2.**
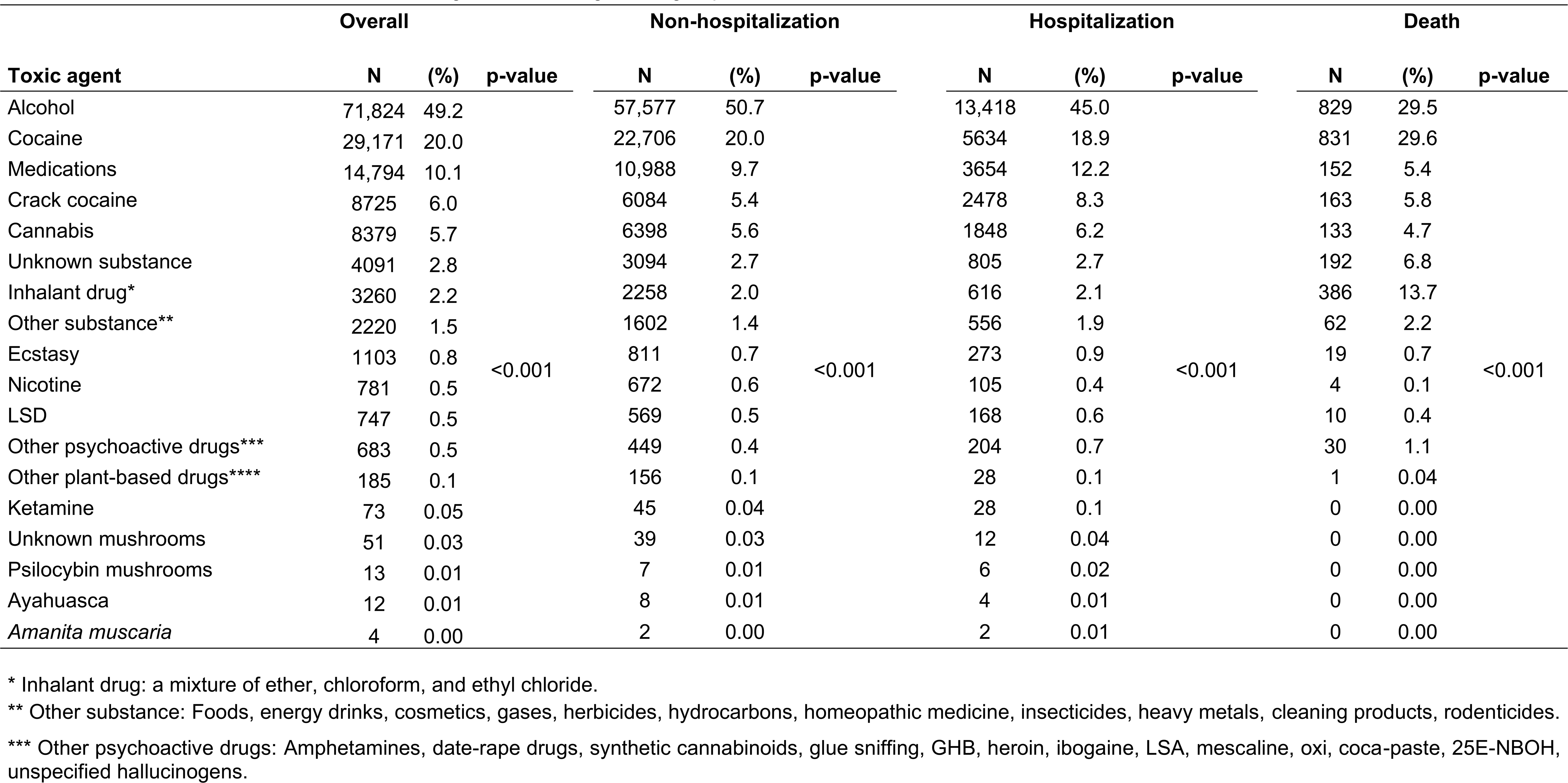
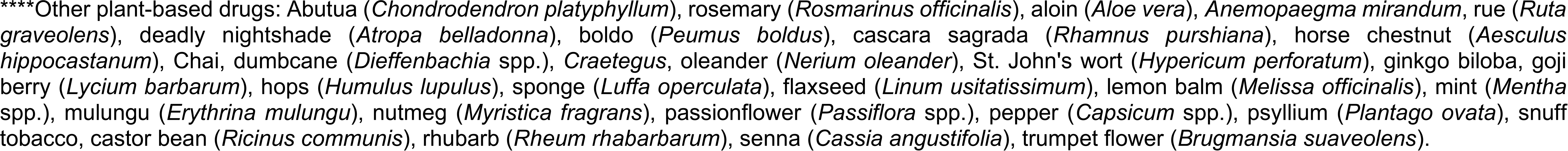
Characteristics and outcomes of toxic agents in the drug abuse group.

#### Demographic characteristics of psilocybin mushroom group

We identified only 13 adverse events, exclusively involving men. Statistically significant variables were the age group 18-25 years (n = 8; 61.5%) (p = 0.006) and the final classification confirmed intoxication (n = 12; 92.3%) (p = 0.002) (Table 1).

#### Demographic characteristics of unknown mushroom group

During the period evaluated, 51 adverse events were reported. Except for co-administration of a toxic agent per case, all other variables presented a statistically significant p-value (p < 0.001). Notably, the majority were men (n = 44; 86.3%), white (n = 29; 56.9%), and individuals aged 18-25 (n = 23; 45.1%). Administration of only unknown mushrooms (n = 34; 66.7%), type of single dose acute exposure (n = 35; 68.6%), and final classification of confirmed intoxication (n = 35; 68.6%) were also evident (Table 1).

#### Outcome data Drug abuse group

##### a. Non-hospitalization

For this outcome (n = 88,493), all variables were statistically significant (p < 0.001) (Table 3). Male individuals (n = 61,744; 69.8%), aged 26-35 (n = 23,162; 26.2%), and white individuals (n = 29,862; 33.8%) stand out. Alcohol (n = 57,577; 50.7%) was the most administered toxic agent (Table 2). There was complete remission in the clinical evolution of 68,351 cases (77.2%) (p < 0.001), followed by partial remission (n = 2952; 3.34%) and lost to follow-up (n = 3823; 4.32%). The outcomes of 13,367 individuals (15.1%) were not recorded.

**Table 3.**
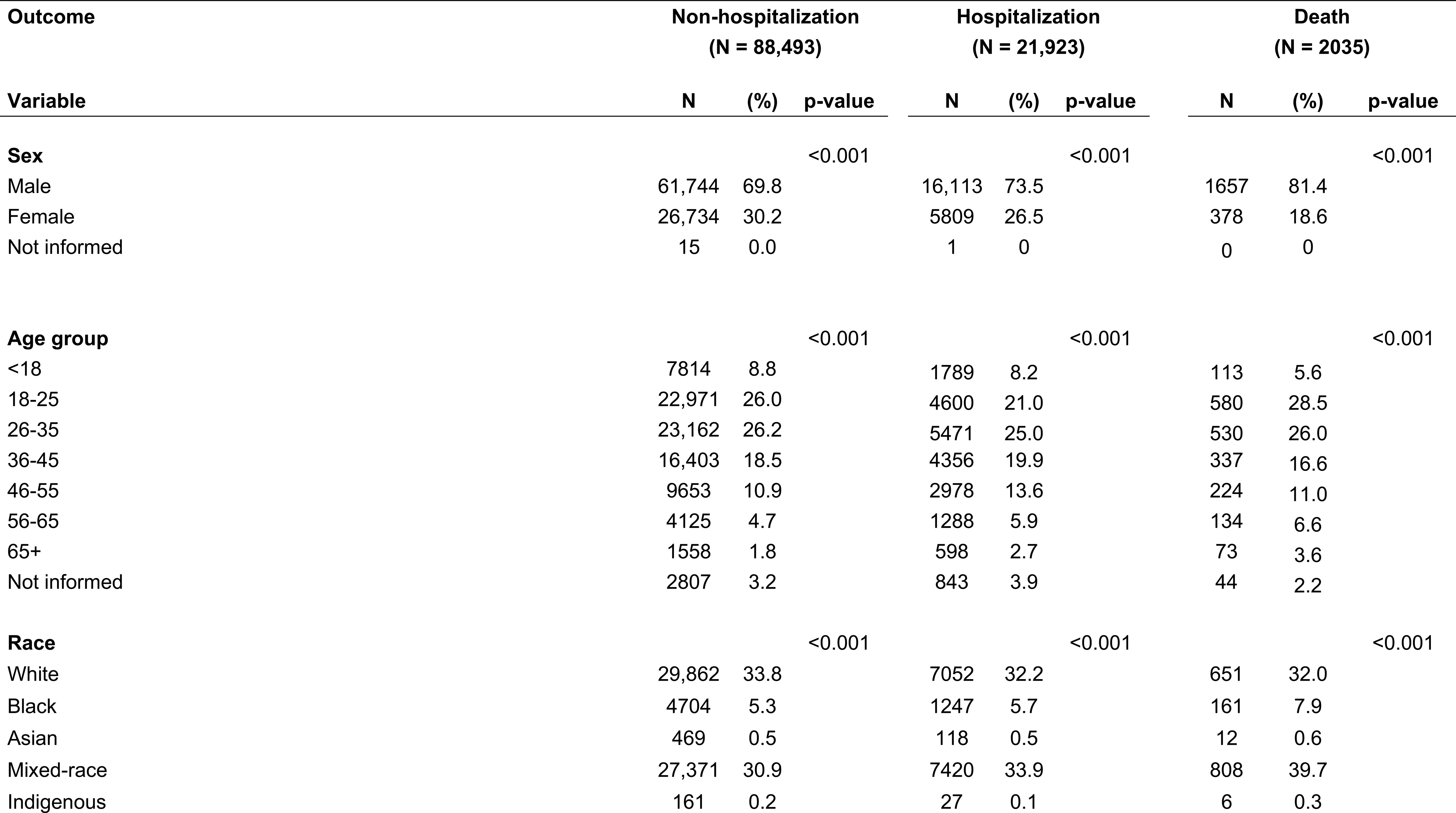
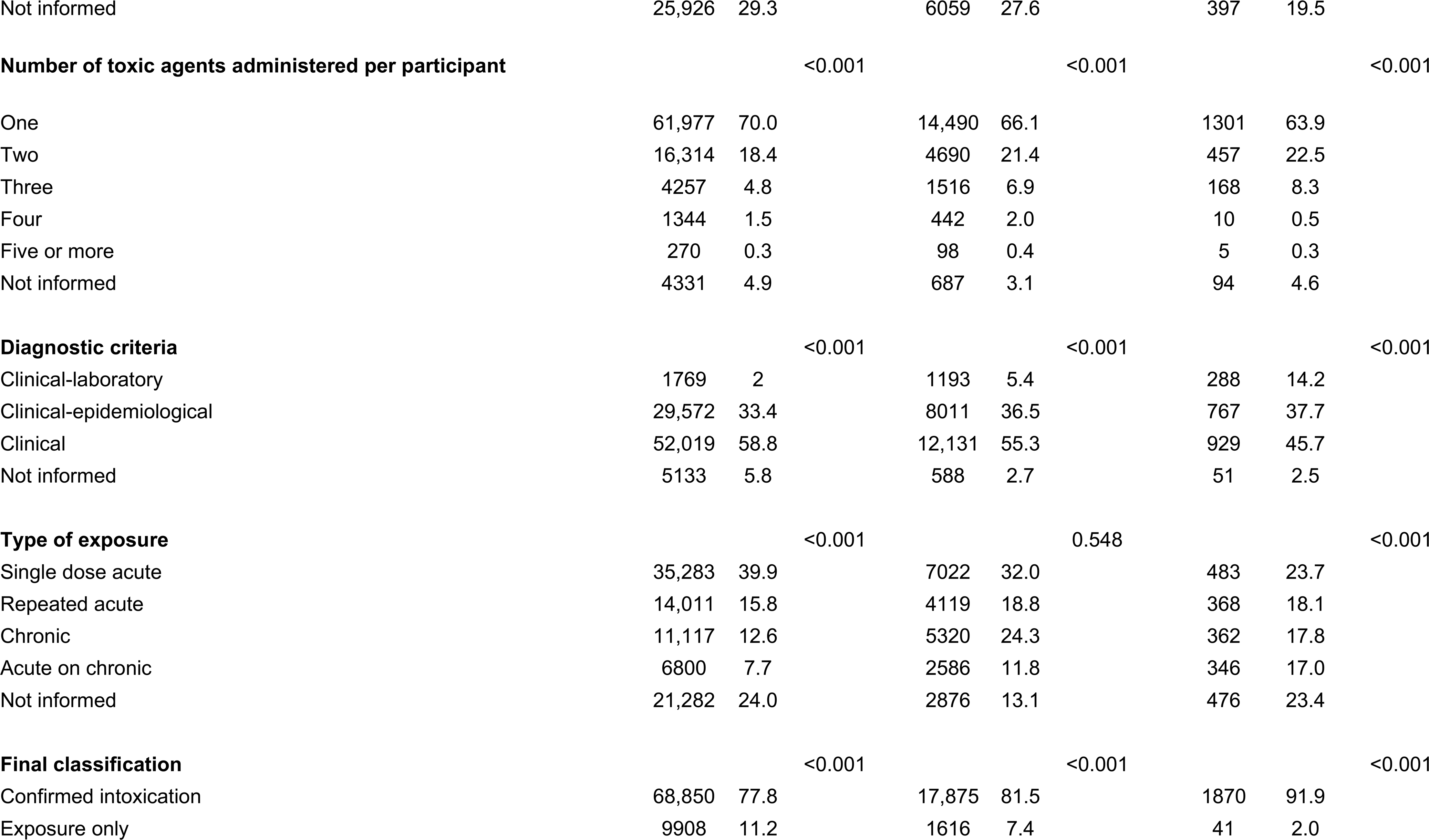
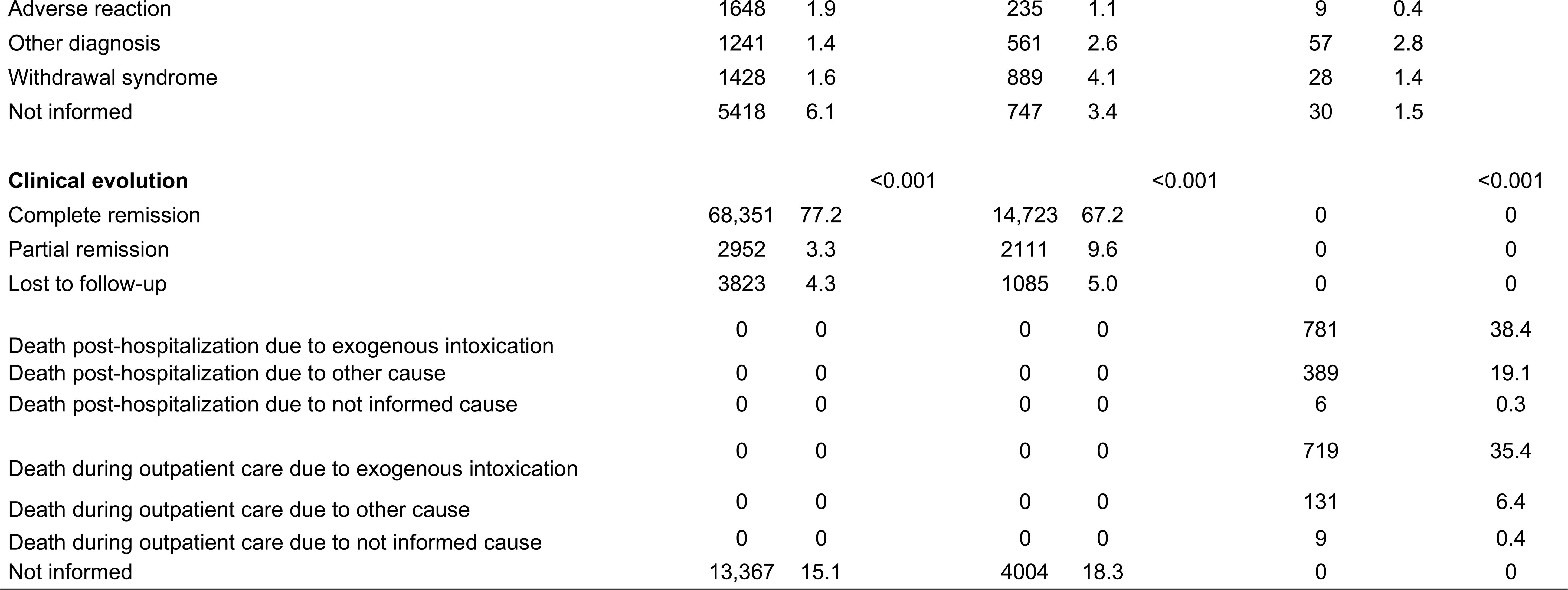
Demographic characteristics by outcomes of the drug abuse group: non-hospitalization, hospitalization, and death.

##### b. Hospitalization

Excluding the type of exposure, the other variables revealed statistically significant associations (p < 0.001) (Table 3). The sample of hospitalized patients (n = 21,923) was mainly composed of men (n = 16,113; 73.5%), mixed race individuals (n = 7420; 33.9%), and those aged 26-35 (n = 5471; 25.0%). Once more, alcohol (n = 13,418; 45.0%) was the main agent observed (Table 2). The record of complete remission corresponded to 14,723 cases (67.2%). Information on the clinical evolution of 4004 hospitalization cases (18.3%) was not recorded.

##### c. Death

There were 2035 deaths recorded between 2007 and 2022. Table 3 shows that all variables demonstrated statistically significant associations (p < 0.001). Men (n = 1657; 81.4%), mixed race individuals (n = 808; 39.7%), and the age group 18-25 (n = 580; 28.5%) were the prominent categories. In contrast to the outcomes of non-hospitalization and hospitalization for drug abuse, cocaine (n = 831; 29.6%) was the major toxic agent (Table 2). Death post-hospitalization due to exogenous intoxication were reported in 781 cases (38.4%), whereas 719 individuals (35.4%) died from exogenous intoxication while receiving outpatient care prior to hospitalization.

#### Psilocybin mushroom group

##### a. Non-hospitalization

None of the variables showed statistical significance. Seven male individuals (100%) were not hospitalized, the majority of whom were white and aged 18-25 years (n = 5; 71.4%). Co-administration of toxic agents occurred in 4 adverse events (57.1%), which included the use of psilocybin mushrooms with alcohol, *Amanita muscaria* (a psychoactive mushroom), cocaine, and lastly, Ecstasy and inhalant drug (a mixture of ether, chloroform, and ethyl chloride). The clinical evolution was favorable in 6 cases (85.7%) (Table 4).

**Table 4.**
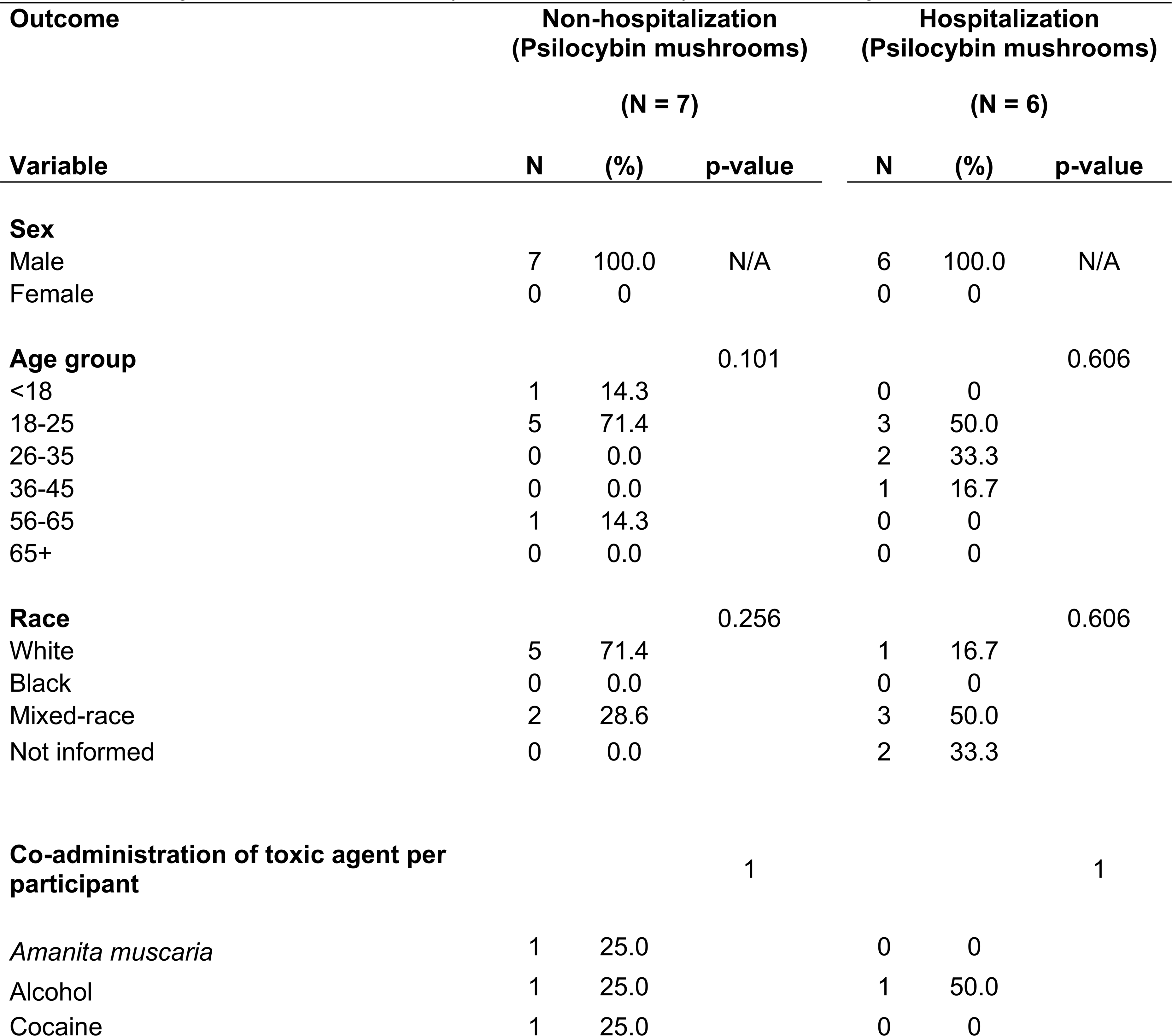
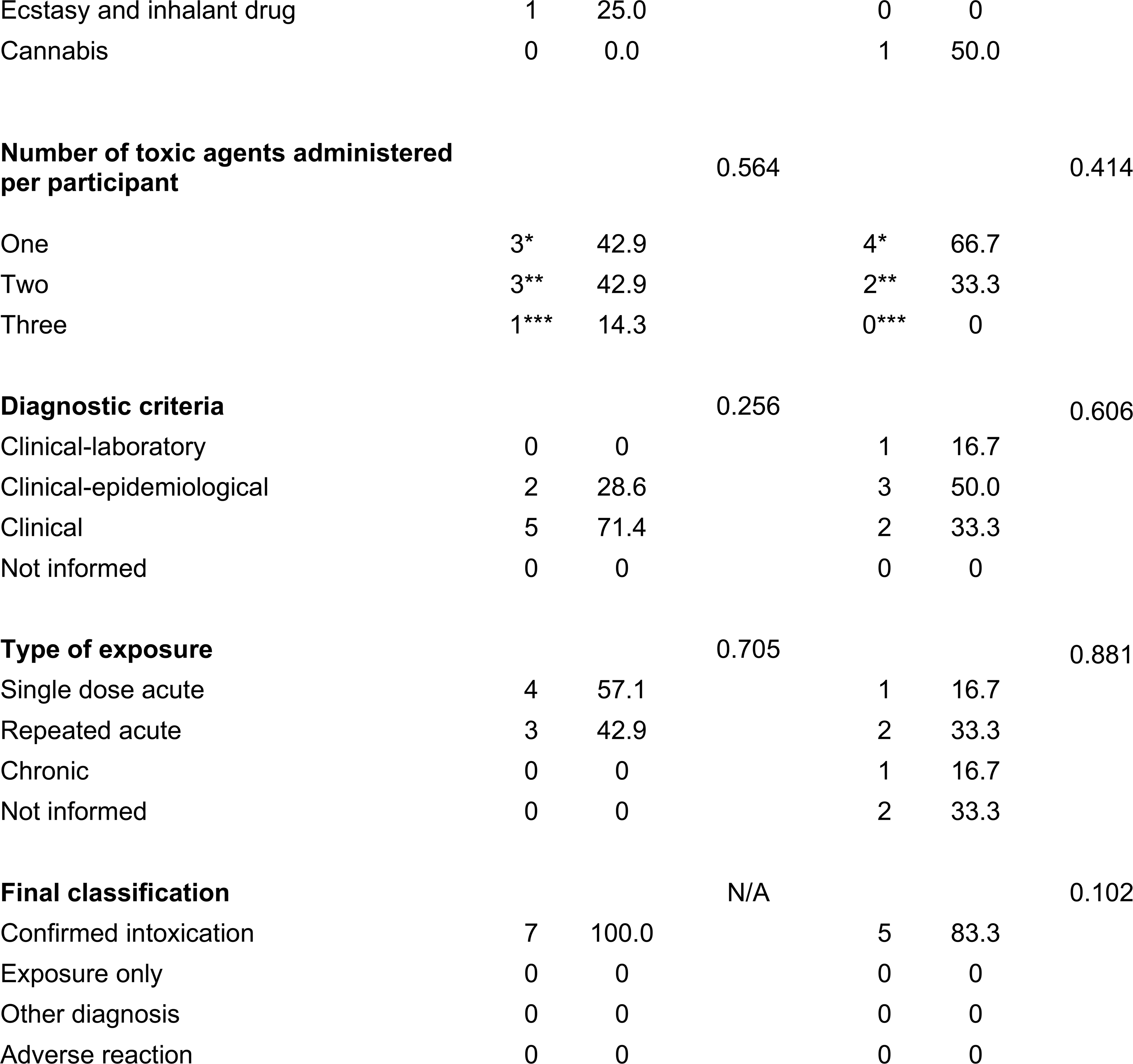
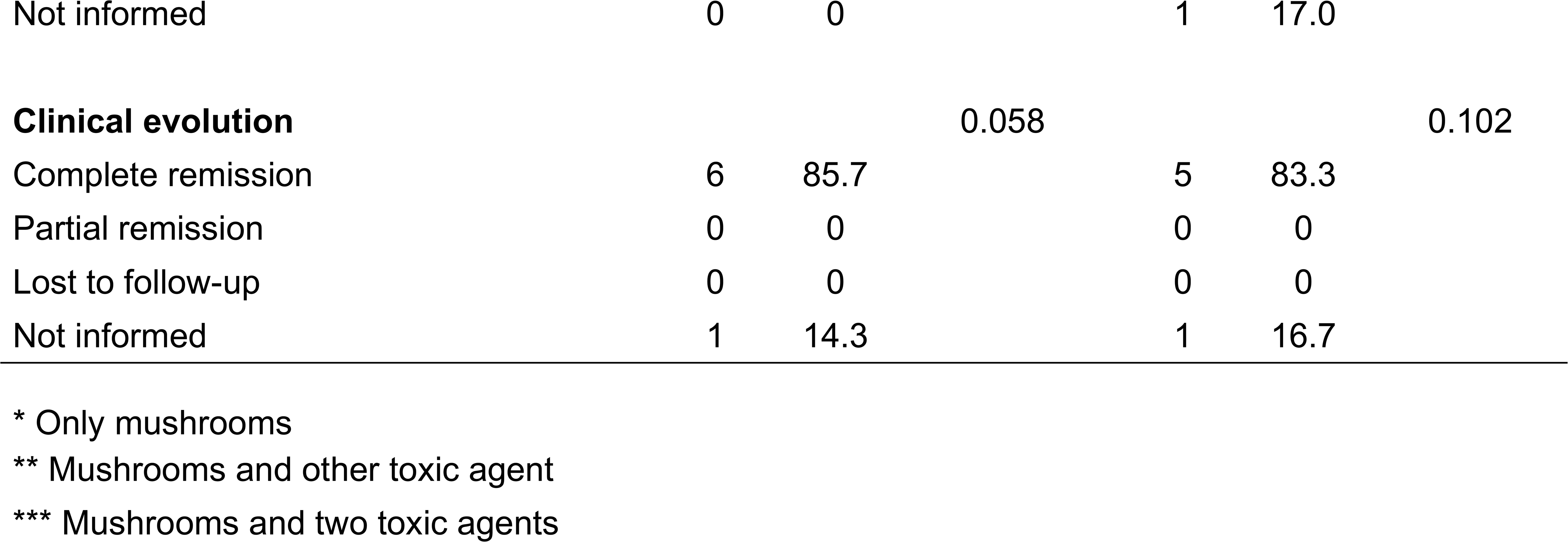
Demographic characteristics by outcomes of psilocybin mushroom group.

##### b. Hospitalization

Hospitalization occurred in 6 cases, all involving men (100%), with a predominance of individuals of mixed race and the age group 18-25 (n = 3; 50%). Administration of psilocybin mushrooms alone corresponded to 4 records (66.7%). Five individuals (83.3%) recovered completely, and only 1 case did not present a clinical outcome. Nonetheless, like the group non-hospitalization for psilocybin mushrooms, none of the variables revealed statistically significant associations (Table 4).

##### c. Death

There were no deaths reported in the psilocybin mushroom group.

#### Unknown mushroom group

##### a. Non-hospitalization

Apart from co-administration of toxic agents, all categorical variables were statistically significant (p < 0.001). Of the 39 non-hospitalizations, 32 (82.1%) involved male individuals aged 18-25 years (n = 18; 46.2%). Ingestion of only unknown mushrooms represented 66.7% of adverse events (n = 26). Overall, the clinical evolution was favorable in 82.1% (n = 32). There was partial remission in 1 case, lost to follow-up in another, and 5 records were not informed (Table 5).

**Table 5.**
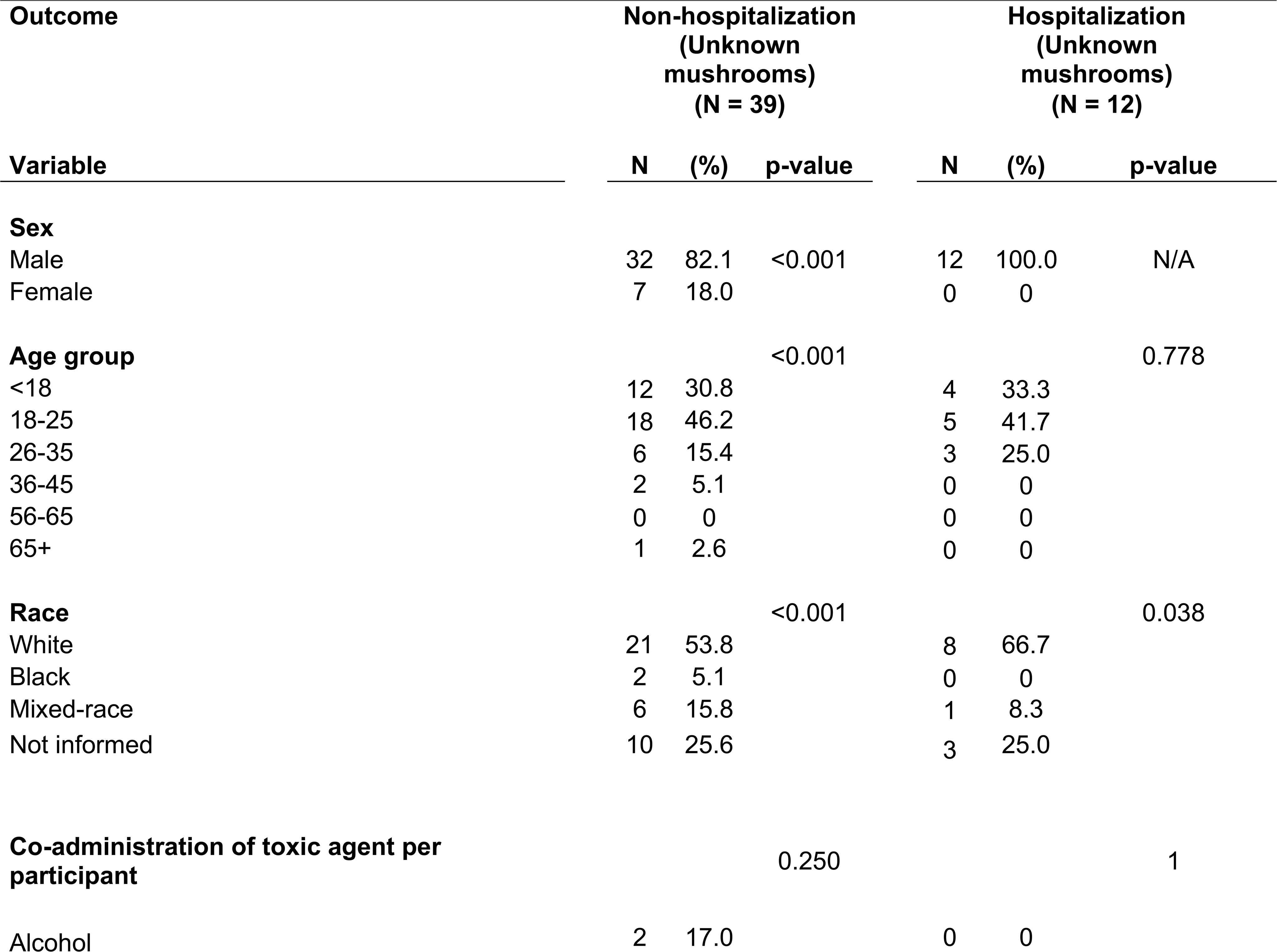
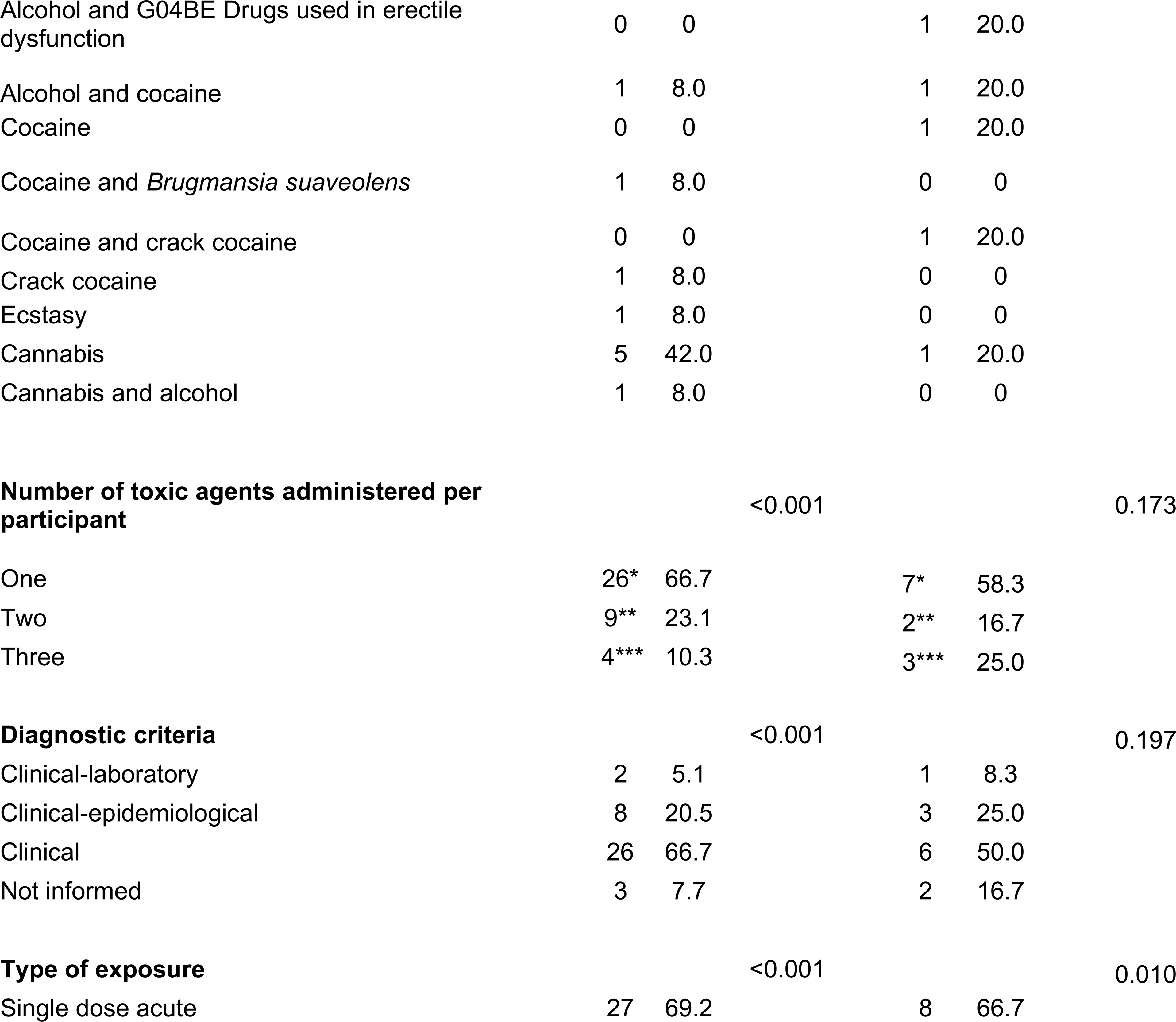
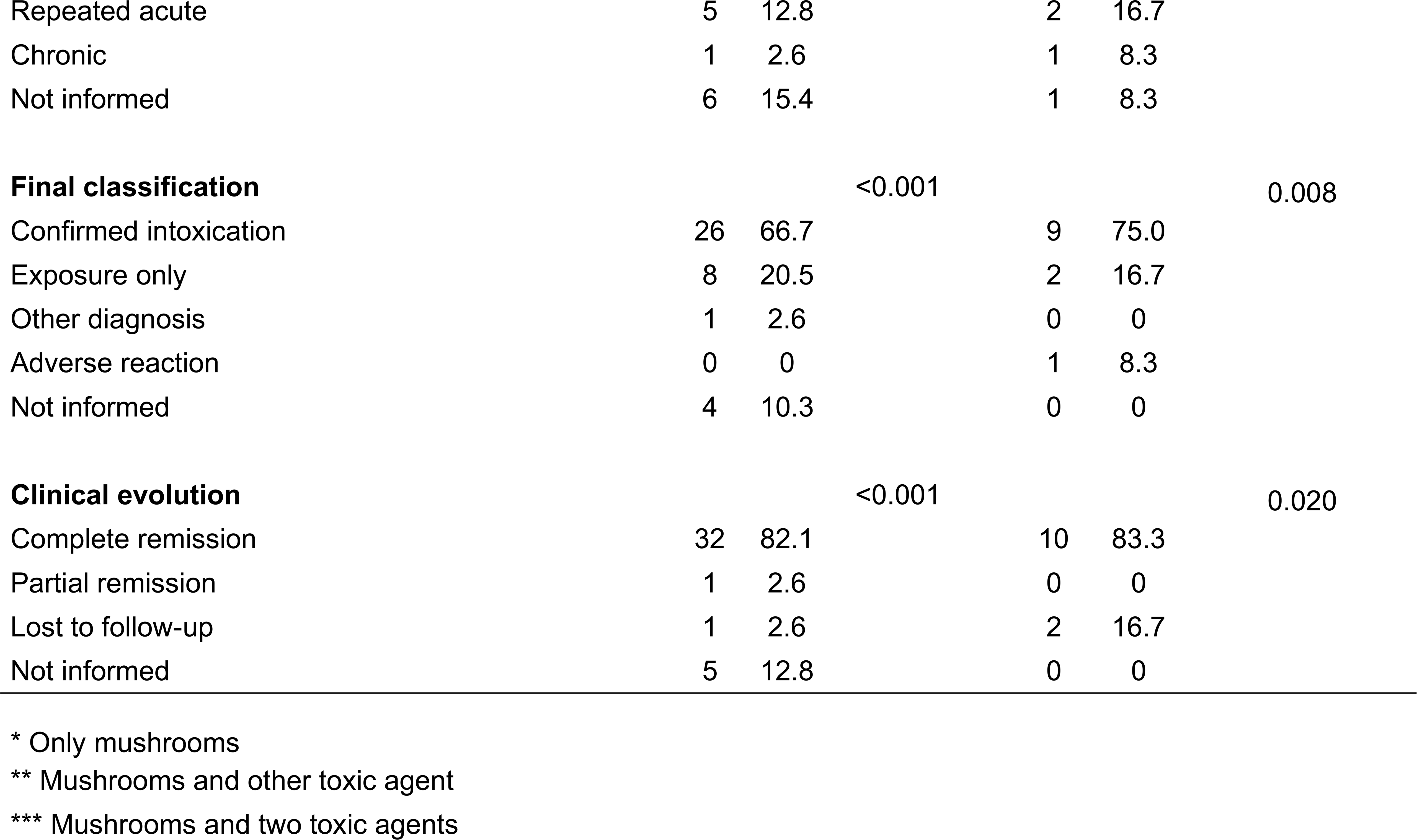
Demographic characteristics by outcomes of unknown mushroom group.

##### b. Hospitalization

Twelve men (100%) were hospitalized for ingesting unknown mushrooms. Only white individuals (n = 8; 66.7%) and those in the age group 18-25 (n = 5; 41.7%) showed a robust statistical association with hospitalization. Additionally, type of exposure single dose acute (n = 8; 66.7%), confirmed intoxication (n = 9; 75.0%), and complete remission (n = 10; 83.3%) also exhibited significant associations (Table 5).

##### c. Death

There were no deaths recorded in the unknown mushroom group.

## Main results

### Prevalence

#### a. Drug abuse group: non-hospitalization, hospitalization and death

In relation to this group, there was a prevalence of 78.7% of incidents that did not result in hospitalization. The prevalence rates of hospitalization and death due to drug abuse were 19.5% and 1.8%, respectively (Table 6).

**Table 6.**
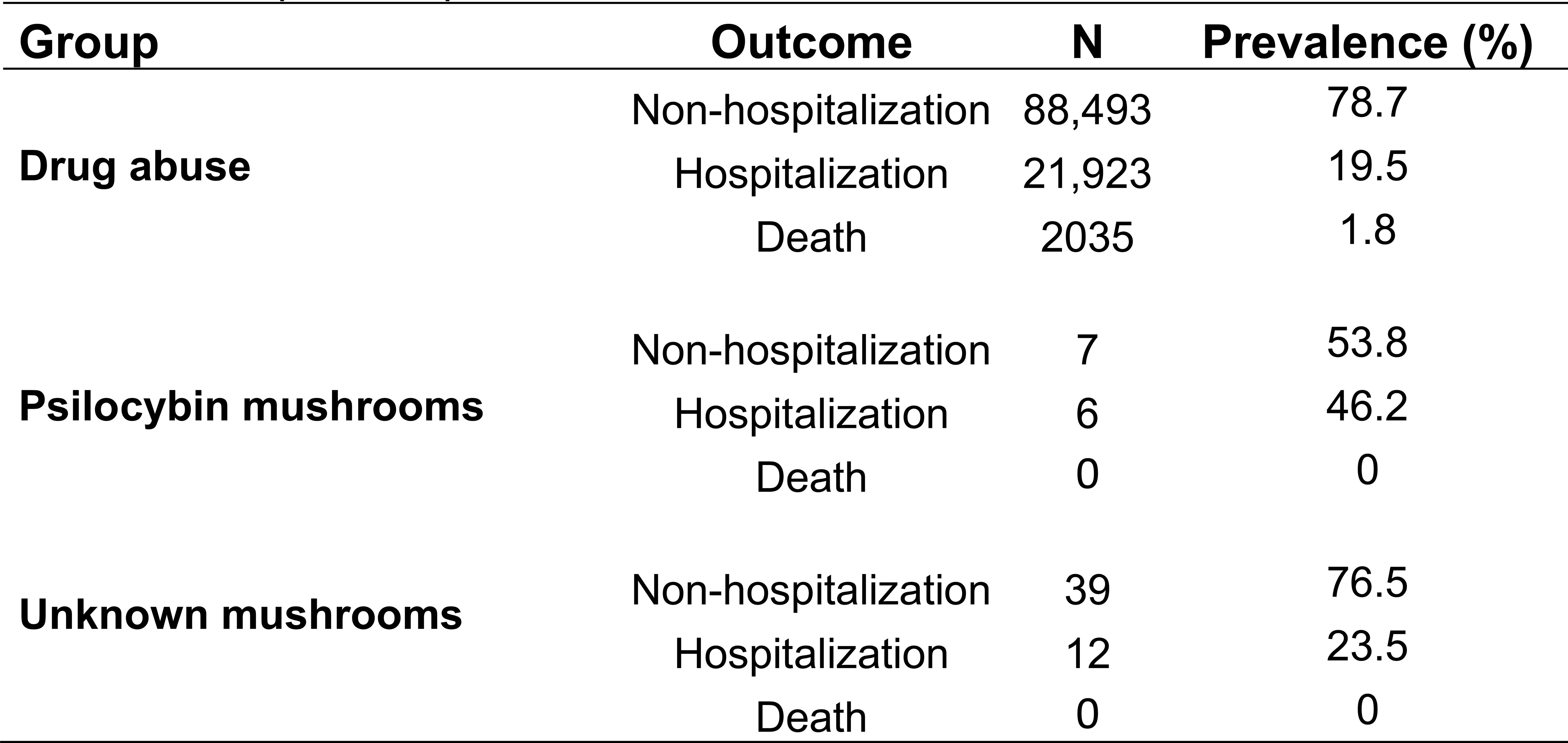
Group-based prevalence of outcomes.

#### b. Preliminary estimates of the prevalence and sample size

Based on the observed preliminary estimates of the prevalence (0.01156%), the proportion formula indicates a minimum sample size of 44,470 individuals who have ever used psilocybin mushrooms to achieve a 95% confidence level with a margin of error of 0.01%. This large sample size helps confirm the low-risk profile of adverse events suggested by the observed prevalence.

#### c. Psilocybin mushroom and unknown mushroom groups: non-hospitalization and hospitalization

The prevalence of non-hospitalization was higher in the unknown mushroom group (76.5%) compared to the psilocybin mushroom group (53.8%) (Table 6). Conversely, hospitalization rates were higher among those consuming psilocybin mushrooms (46.2%; 99% CI: 10.6% - 81.6%) compared to the unknown mushroom group (23.5%; 99% CI: 8.3% - 38.7%). Despite this trend towards higher hospitalization rates in the psilocybin mushroom group, the difference was not statistically significant.

#### d. Drug abuse group: outcomes rates by toxic agent

To illustrate the impact of toxic agents, Figure 2 estimates hospitalization, non-hospitalization and death rates due to exogenous poisoning and from other causes. We can see that psilocybin mushrooms represented 0.02% of all hospitalizations, while the rate for unknown mushrooms is slightly higher, 0.04%. Most hospitalizations cases involved alcohol (45.0%), cocaine (18.9%), medications (12.2%), crack cocaine (8.3%) and cannabis (6.2%). Deaths due to exogenous intoxication were represented mainly by cocaine (33.3%) and, from another cause, by alcohol (51.9%).

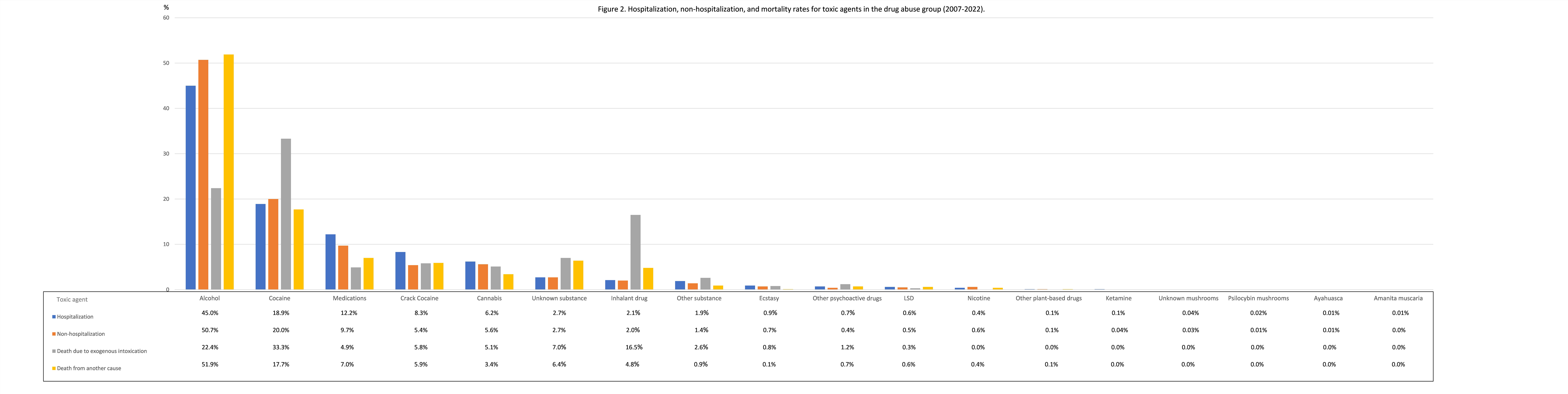

### Other analyzes

#### Prevalence of toxic agent-associated outcomes in different exposure groups

A descriptive analysis of hospitalization and mortality rates across various toxic agent exposure groups revealed significant insights (Table 7). Notably, inhalant drug had the highest mortality rate (11.8%), followed by unknown substance (4.7%) and other psychoactive drugs (4.4%). For hospitalization, *Amanita muscaria* stood out with a high rate (50%), though the sample size was small. Additionally, ketamine showed a high hospitalization rate (38.4%) despite no reported deaths. Common substances like alcohol (18.7% hospitalization) and cannabis (22.1% hospitalization) displayed moderate hospitalization rates, while cocaine (19.3% hospitalization) and crack cocaine (28.4% hospitalization) presented higher risks. Medications (24.7% hospitalization) and ecstasy (24.8% hospitalization) also showed notable rates. While causality cannot be established, these findings highlight trends in hospitalization and mortality outcomes across exposure groups.

**Table 7.**
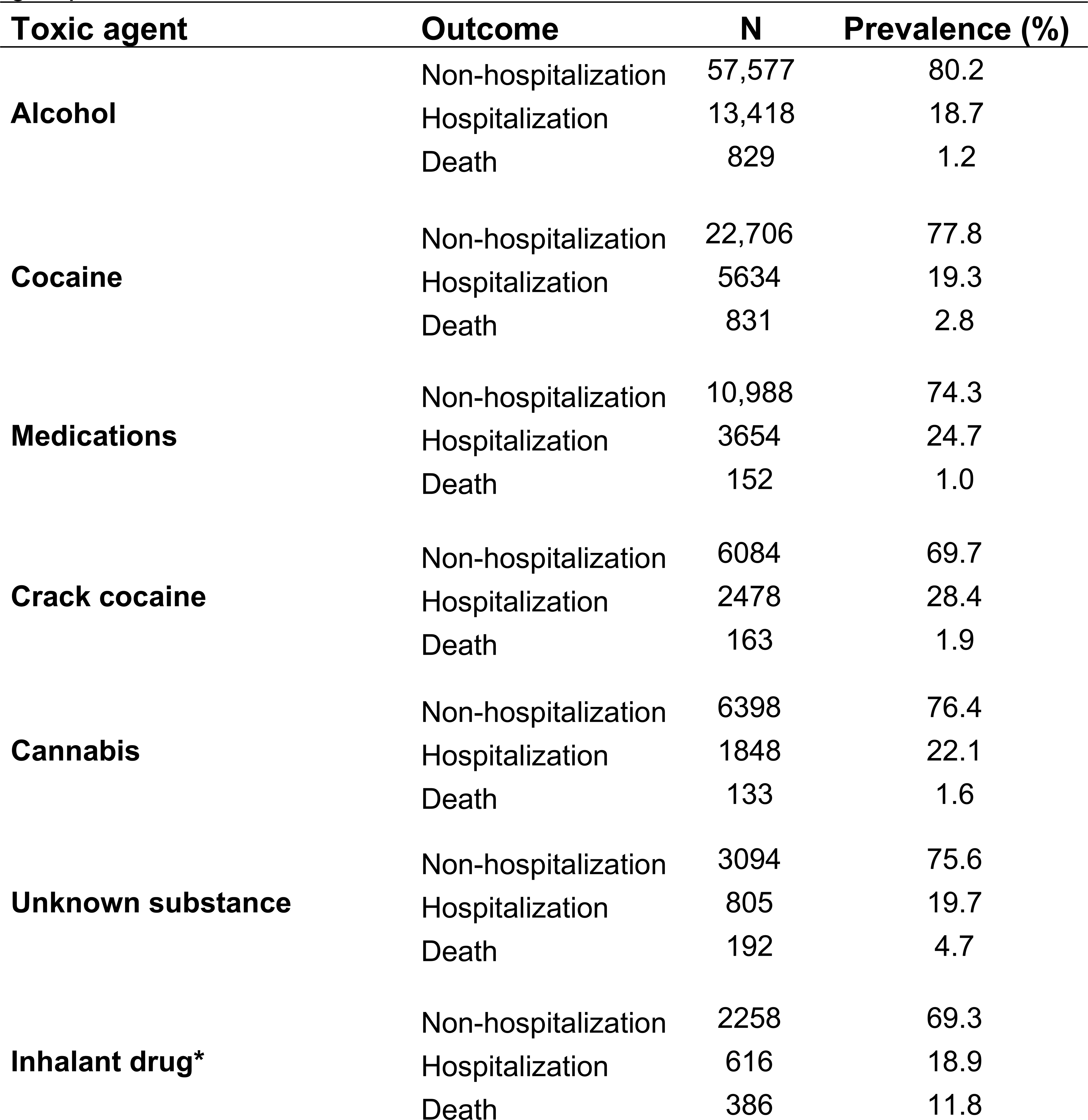
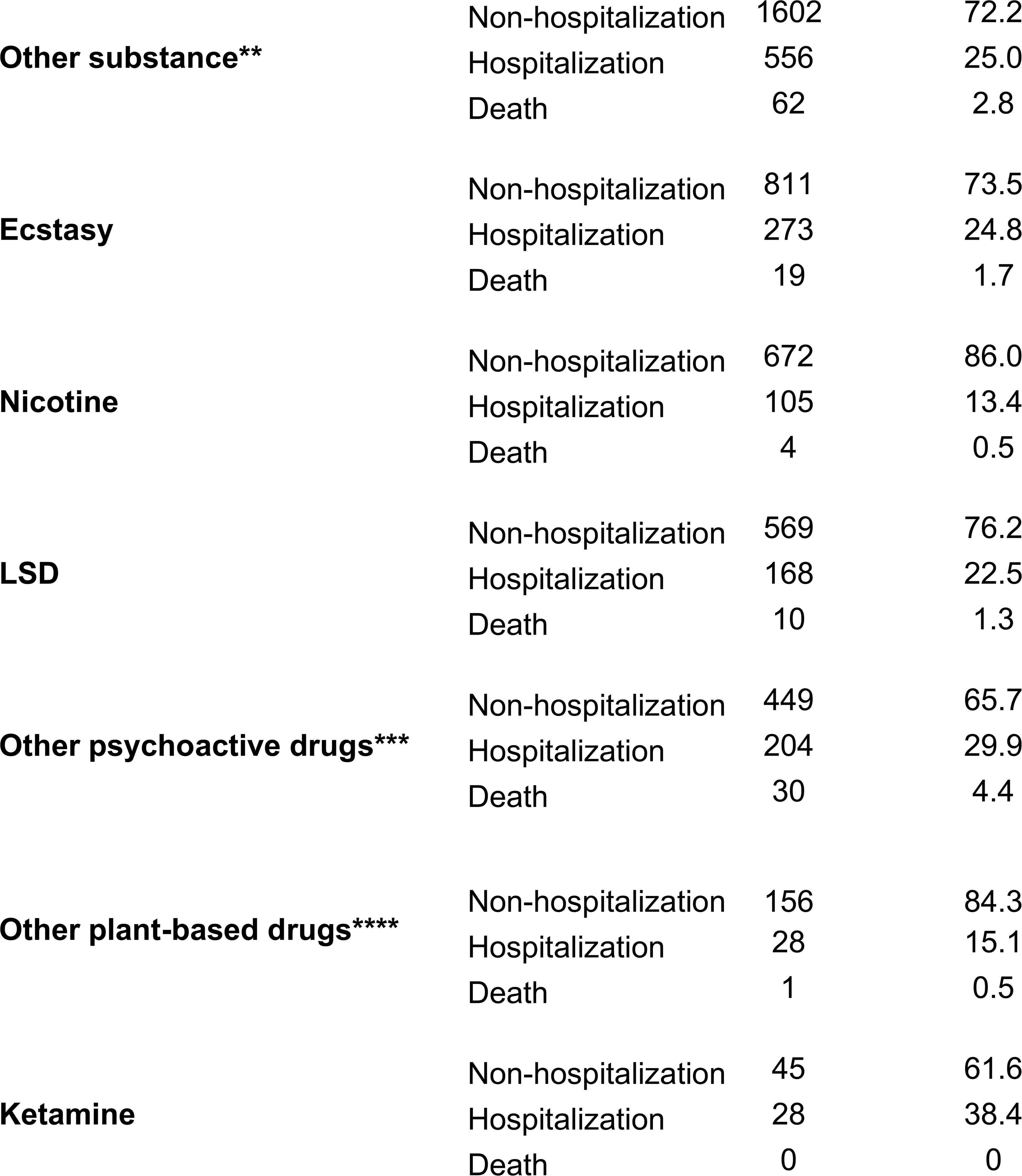
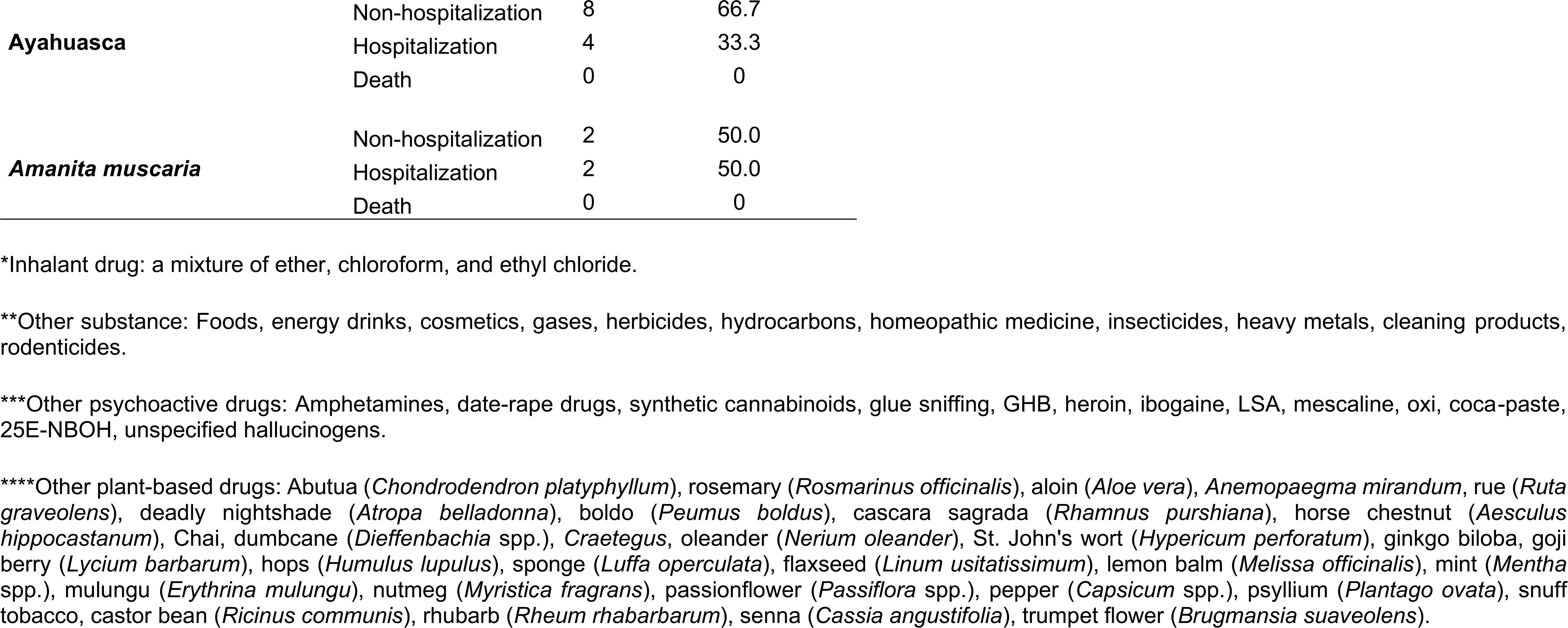
Prevalence of toxic agent-associated outcomes in different exposure groups.

## Discussion

This is the first study to present the adverse event profile associated with drug abuse in Brazil. We reveal that psilocybin mushrooms represent a low-risk adverse event compared to other toxic agents frequently abused by the Brazilian population. Our findings raise questions about the scientific incoherence of current drug policy (BRASIL, 1998; Levin et al., 2022) and point out that psilocybin mushrooms do not pose a significant risk to Brazilian public health and do not have abuse potential.

Further supporting the low-risk profile of psilocybin mushrooms, data from the Global Drug Survey (Winstock A et al., 2017) highlight their security aspects. Among the over 12,000 individuals who reported using psilocybin mushrooms in 2016, only nineteen (0.2%) needed medical attention (Kopra et al., 2022; Winstock A et al., 2017). Of these, eight (42%) were hospitalized but promptly recovered within twenty-four hours. From 2007 to 2022, the prevalence of hospitalization involving psilocybin mushrooms in Brazil was 46.2% (Table 6), a rate comparable to annual data from the Global Drug Survey. While this rate appears higher compared to hospitalizations observed for other toxic agents (Table 7), caution is warranted when interpreting these data due to the extremely small sample size of adverse events recorded in the historical series, which may lead to an overestimation of the true hospitalization rate.

Understanding psilocybin’s safety profile is crucial, especially considering the hospitalization rate in Brazil. Although we have not identified the symptoms of adverse events leading to hospitalization, psilocybin is generally considered tolerable and safe, even in a variety of psychiatric and substance dependence conditions (Kaminski & Reinert, 2023). The most common acute side effects are transient, resolving within forty-eight hours, and include nausea, headaches, increased blood pressure, and heart rate (Kaminski & Reinert, 2023; Yerubandi et al., 2024). Furthermore, variations in psilocybin potency across species, strains, growth, and storage conditions make it difficult to estimate dosage (Gable, 2004). This significantly increases the risk of challenging experiences and adverse reactions like anxiety, paranoia, and panic attacks, which may require medical attention. (Kopra et al., 2022, 2023).

Regarding psilocybin and unknown mushrooms incidents, our analysis also explored demographic characteristics associated with adverse events. Interestingly, we found a statistically significant skew towards males within the eighteen to twenty-five age group who experienced adverse events. On the other hand, a systematic review demonstrated that gender may not be a factor in psychedelic-related incidents (Aday et al., 2021). This aligns with Kopra et al., 2022, who highlight factors like inadequate “set” and “setting,” younger age, and mixing substances as key contributors to adverse psychedelic experiences. Other exploratory study suggested a link between younger age and the intensity of challenging psychedelic experiences (Ko et al., 2023), which may necessitate urgent medical care (Kopra et al., 2022). Consistently, our findings correspond with the results of this previous research. The underrepresentation of older age groups in our data may be due to the influence of biological or psychological factors associated with aging. These factors could include the development of stronger coping mechanisms to manage negative or challenging psychedelic experiences, as suggested by Ko et al., 2023. For instance, older adults may have developed a greater sense of emotional stability, which can mitigate the intensity of challenging psychedelic experiences and reduce the likelihood of seeking medical attention.

Our findings on demographics and risk factors also underscore the need for more robust data on psilocybin mushroom consumption patterns in Brazil. Although a comprehensive dataset was unavailable, we conducted a sample size calculation. The result indicated that 44,470 individuals would be required to achieve the desired level of accuracy. This minimum sample size is essential to ensure that the preliminary estimates of prevalence fall within the specified margin of error, thereby providing reliable results for a low-risk adverse event profile in the population. Given the unrestricted sales since the early 1990s and the widespread availability of psilocybin mushrooms in Brazil’s biome, it is plausible that a significant number of individuals have used them historically without requiring medical attention. This plausibility is further supported by the favorable pharmacological safety profile of psilocybin mushrooms, which may account for the rarity of such adverse events (Johnson et al., 2018; Jones et al., 2023; Kaminski & Reinert, 2023). Nevertheless, this interpretation should be approached with caution, emphasizing the importance of obtaining consumption pattern data in the future to validate these assumptions.

Due to the lack of self-reported data in Brazil, exploring other drug safety profile findings can provide valuable insights. Like the Global Drug Survey results (Winstock A et al., 2017), Table 2 and Figure 2 indicate that psilocybin mushrooms are physiologically safe and represent a low-risk adverse event profile compared to other toxic agents such as alcohol, cocaine, crack cocaine, medications and cannabis. Additionally, a study by the National Survey on Drug Use and Health reinforced these findings by showing that psilocybin use is not associated with criteria for abuse and dependence, such as tolerance, physical/emotional problems, and significant personal issues (Jones et al., 2023).

In contrast, our analysis did not identify any fatalities associated with psilocybin mushrooms or unknown mushrooms, unlike the 1.8% mortality rate observed in the drug abuse group (Table 6). Notably, this drug abuse group predominantly consists of younger men (Table 3) and individuals using cocaine (Table 2, Figure 2). Among various toxic agent exposure groups, inhalant drug showed the highest mortality rate at 11.8%. Nevertheless, scientific literature reports only rare instances of fatalities associated with psilocybin mushroom use in specific situations.

For instance, one case involved the ingestion of *Psilocybe semilanceata* (a wild mushroom) by a twenty-two-year-old man, who experienced nausea and vomiting, followed by loss of consciousness, coma, and death (Gerault A & Picart D, 1996). Another case concerns a twenty-four-year-old woman, a heart transplant recipient, who ingested an unknown quantity of psilocybin mushrooms and was admitted to the hospital after a cardiac arrest that resulted in death. The plasma toxicology test revealed the presence of psilocin (the active metabolite of psilocybin) and THC (tetrahydrocannabinol) (Lim et al., 2012), which highlights the co-administration of substances. Between 2000 and 2023, in Australia, ten deaths associated with psilocybin were reported, mainly among young men around twenty years old. The most common circumstance of death was a traumatic accident (n = 4), followed by an undetermined cause (n = 4) and multiple drug intoxication (n = 2) (Darke et al., 2024). These cases could have been avoided in a regulatory scenario. Regulation is crucial to educate the population and prevent serious adverse events, including those from ingesting wild mushrooms, mixing with other psychoactive drugs, or using them in dangerous settings.

Even so, it’s important to note that deaths caused by psilocybin mushroom poisoning are extremely rare. This rarity might be due to the lack of evidence for psilocybin causing brain damage, organ failure, or addiction (Johnson et al., 2018; Jones et al., 2023; Kaminski & Reinert, 2023). Additionally, the amount of psilocybin needed for a fatal overdose is estimated to be around six grams, which is equivalent to roughly ten kilograms of fresh mushrooms. Because of this high amount, accidental overdose by eating psilocybin mushrooms is highly unlikely as vomiting would likely occur before a dangerous amount is absorbed (Johnson et al., 2018).

While psilocybin mushrooms themselves appear to have a low-risk profile, our analysis also identified fifty-one adverse events associated with unknown mushrooms. Brazil’s diverse biome (Plazas & Faraone, 2023) and the common practice of foraging for fresh mushrooms complicate the confirmation of the type of fungi involved — whether they were toxic, contained psilocybin, or had other psychoactive properties. Notably, a 1.97-fold increase in prevalence ratio was observed among those hospitalized in the psilocybin mushroom group compared to the unknown mushroom group. However, the overlap of the confidence intervals suggests the difference in hospitalization prevalence between the two groups is not statistically significant, which could be attributable to chance and may not represent a true discrepancy. A possible explanation for the adverse events in the unknown mushroom group is misidentification, with individuals potentially mistaking toxic or non-psychoactive mushrooms for psilocybin mushrooms. This context highlights the importance of regulation, as it would prevent serious adverse events by facilitating access to psilocybin mushrooms grown in a controlled environment.

The potential dangers of unknown mushrooms underscore the importance of considering psilocybin’s therapeutic potential and the current trend of self-treatment. Although our study focused on adverse events related to drug abuse, the increasing trend of self-treatment with psilocybin mushrooms (Kopra et al., 2023) cannot be overlooked. This trend is driven by growing public interest in the mounting scientific evidence for psilocybin’s benefits (Goodwin et al., 2022, 2023; Griffiths et al., 2016; Irizarry et al., 2022), such as long-term improvements in anxiety and depression symptoms, reduced alcohol consumption, and others (Nayak et al., 2023). The slow development of effective mental health medications (Nutt et al., 2024), inconsistent regulations (BRASIL, 1998; Levin et al., 2022), and a lack of psilocybin knowledge among healthcare professionals (Meyer et al., 2022), all contribute to people seeking self-treatment. Consequently, many turn to personal experimentation rather than professional guidance. A survey that examines information-seeking behavior of people using psychedelics shows that the most common source of participants’ information was their own experimentation (Kruger et al., 2023). Understanding these factors influencing self-treatment is essential for contextualizing the adverse event profile of psilocybin mushrooms observed in our analysis.

Although the trend of self-treatment with psilocybin mushrooms highlights the potential risks associated with unsupervised use, a systematic review with meta-analysis suggests that psychedelics are generally well-tolerated with a low likelihood of serious adverse events when administered in a controlled environment with appropriate screening criteria (Romeo et al., 2024). This highlights the importance of preparation and harm reduction. Therefore, we agree with the idea that psilocybin mushrooms administration in a regulated scenario should be preceded by proper guidance (Kruger et al., 2023) and conducted in suitable settings. Strategies to achieve this goal may include ongoing education for healthcare professionals (Meyer et al., 2022), sharing information with both patients and people who use psilocybin mushrooms for spiritual purposes (Kruger et al., 2023), and exploring the motivations behind psychedelic use (Basedow & Kuitunen-Paul, 2022). Even with these measures in place, it’s important to acknowledge that psilocybin, like any psychoactive substance, is not entirely risk-free.

Potential harms from psilocybin include dangerous behavior in unprepared and unsupervised individuals, as well as exacerbation of mental illness in those with or predisposed to psychotic disorders (Johnson et al., 2018). Contraindications to psilocybin ingestion include pregnancy, breastfeeding, personal or family history of schizophrenia, personality disorders, self-harm, suicidal ideation, epilepsy, and cardiovascular diseases (Johnson et al., 2018). These contraindications underscore the necessity for thorough screening and risk assessment before considering psilocybin administration. Additionally, the potential for interaction with other substances or medications should not be overlooked. Psilocybin can interact with a range of substances, including antidepressants, lithium, and other medications, potentially enhancing or diminishing effects and leading to unpredictable and possibly harmful outcomes (Harris, 2023; Thomann et al., 2024). All potential risks are heightened in unregulated environments where users may not be aware of these interactions.

Despite this, our survey confirms that psilocybin uses, and associated harms are lower compared to typically abused drugs (Johnson et al., 2018; Winstock A et al., 2017). In a regulated scenario, clear communication regarding precautions and potential risks becomes crucial to avoid adverse events. Continuous educational programs can enable both patients and individuals seeking spiritual experiences with psilocybin to make informed choices, potentially fostering safer and more positive outcomes.

Since the previous section highlighted the potential risks associated with psilocybin use, a well-regulated environment becomes crucial. While synthetic psilocybin has shown promise in treating various medical conditions, particularly depression, research has primarily focused on this form (Irizarry et al., 2022). However, preclinical evidence (Hernandez-Leon et al., 2024) and observational studies (Kopra et al., 2023; Nayak et al., 2023) also suggest benefits of *Psilocybe cubensis*, prompting new clinical research underway (Escamilla et al., 2023). Moreover, evidence indicates that these fungi may have potent and long-lasting therapeutic effects compared to synthetic psilocybin (Shahar et al., 2024). This highlights the urgent need for regulatory oversight, as increasing self-medication (Australian Institute of Health and Welfare, 2020; Kopra et al., 2023), may lead to missing crucial information on its efficacy and safety.

Another aspect to consider is the legal and social implications of psilocybin mushroom use. Motivated by the growing public interest in psychedelics (Australian Institute of Health and Welfare, 2020; Kopra et al., 2023), we hope this study contributes to an evidence base that will inform future regulatory policies. Currently, in our country, there is no regulation on the sale, possession, or cultivation of psilocybin mushrooms, resulting in unrestricted trade, arbitrary arrests, and an increasing number of individuals resorting to self-treatment with these fungi (BBC News Brasil, 2023; UOL, 2023), which are also freely available in nature (Plazas & Faraone, 2023).

Considering this situation, our study, along with advances in the reclassification of psilocybin in Australia (Haridy, 2023) and Oregon (USA) (Holoyda B, 2023), encourages regulatory discussions in Brazil. Regulation may prevent arbitrary arrests and adverse events by allowing for the reception and education of patients seeking therapeutic benefits. Under the clinical logic of regulatory agencies, psilocybin mushroom regulation could structure long-term data collection, enabling the implementation of a pharmacovigilance system. This system would complement the findings of controlled studies with synthetic psilocybin, contributing to a better understanding of its efficacy and safety.

Conversely, as psychedelic research gains global importance, it is essential to respect indigenous communities by recognizing their intellectual property and avoiding cultural appropriation in a regulatory environment (Celidwen et al., 2023; Omágua-Kambeba et al., 2023) We must consider that the ceremonial use of psilocybin precedes its clinical use, and that this substance has been used for centuries in religious or shamanic contexts (Carod-Artal, 2015; Guerra-Doce, 2015; Nayak et al., 2024; Rucker & Young, 2021). Hence, inviting indigenous communities to participate in regulatory deliberations is vital to ensure their perspectives and rights are respected. However, there are still no established ethical principles regarding the spiritual practices involving psilocybin mushrooms in Brazil, highlighting the need for further research in this area. In view of this, we propose that the anthropological issues related to psilocybin mushrooms, including existing spiritual practices (Rucker & Young, 2021), be initially examined under the Brazilian constitutional guarantee of freedom of worship. This approach was similarly applied in the regulatory process for ayahuasca in our country (Labate & Feeney, 2012), which also demonstrated a low-risk adverse event profile, similar to these fungi (Figure 2) (Table 7).

Finally, in light of the findings and key points discussed in this study, a call is made for ANVISA and the National Drug Policy Council (CONAD) to engage in evidence-based regulatory discussions. It is imperative to address the legal implications surrounding psilocybin mushrooms in Brazil. The ambiguous legal status of these fungi creates uncertainty and barriers to safe access, hindering both research efforts and the implementation of harm reduction strategies. Clarifying and updating existing laws and regulations to align with evolving scientific evidence and societal perspectives is crucial for establishing a coherent regulatory framework that prevents arbitrary arrests and ensures availability of psilocybin mushrooms for both therapeutic and ceremonial purposes. Examples of regulatory models, such as those implemented in Australia and Oregon, can serve as references for creating effective regulations in Brazil.

Other considerations for evidence-based regulatory discussions could suggest:

1. Education and training

- Implement continuing education programs for healthcare providers on psilocybin mushroom use. Ensure that training encompasses both clinical and ceremonial settings.
- Facilitate open communication between healthcare providers and patients.
- Engage with spiritual communities to provide information and clinical support regarding ceremonial use of psilocybin mushrooms.
2. Harm reduction strategies in self-treatment contexts

- Implement public campaigns providing comprehensive information materials to educate the population about psilocybin mushrooms, its potential benefits, risks, and responsible use, promoting informed decision-making.
- Emphasize the importance of preparation, risk assessment, as well as safe and supportive settings for psilocybin mushroom use, to minimize adverse events.
- Educate the public about the potential dangers of mixing psilocybin mushrooms with other substances, including medications and psychoactive drugs.
- Develop and disseminate information on the risks associated with wild mushroom ingestion, including identification, potential contaminants, and adverse effects.
3. Indigenous community inclusion

- Involve indigenous people in regulatory discussions, considering potential ceremonial use of psilocybin mushrooms by local indigenous communities, as well as in research aimed at establishing ethical principles regarding spiritual practices involving these fungi.
- Respect indigenous knowledge, practices, and intellectual property rights related to psilocybin mushrooms and other entheogens, ensuring that regulation frameworks are developed with sensitivity to indigenous cultural practices and beliefs.
- Avoid cultural appropriation by collaborating with indigenous communities as equal partners in research and decision-making.
4. Pharmacovigilance system

- Implement a robust pharmacovigilance system to monitor the safety and efficacy of psilocybin mushroom use in clinical, ceremonial, and self-treatment settings, potentially linking it to stores that sell these fungi.
- Collect and analyze data on adverse events, long-term effects, and overall outcomes associated with psilocybin mushroom use, to complement controlled clinical trials and provide a broader understanding of psilocybin’s safety and efficacy profile.
5. Clinical research in Brazil

- Encourage and support well-designed clinical research on psilocybin mushrooms in Brazil to generate local evidence and inform regulatory decisions.

## Limitations

This retrospective cross-sectional study provides a snapshot of historical data, but it cannot establish causal relationships. The results do not necessarily indicate that a specific toxic agent caused hospitalization and/or death at the individual level. Furthermore, we did not analyze the associations between variables and groups of toxic agents in relation to the observed outcomes. Our objective was to evaluate the profile of adverse events associated with drug abuse, with emphasis on psilocybin mushrooms and unknown mushrooms. In observational studies, confidence intervals (CIs) are commonly used to assess data variability. However, in instances where the outcome is exceptionally rare and the sample size is small or very limited, calculating the CI may not contribute significantly to the analysis. This is because the CI can be excessively wide, which complicates precise interpretations by encompassing both positive and negative variability. In such cases, a very wide CI relative to a small interval may obscure rather than clarify insights.

Another significant problem is the lack of identification of psilocybin mushroom species, which can introduce variability and uncertainty into results, making hospitalizations difficult to interpret. Nevertheless, the widespread availability of *Psilocybe cubensis* in both specialized companies and the Brazilian biome suggests that this species is likely involved in most cases.

We also did not examine all circumstances of adverse events involving psilocybin mushrooms for clarity. From the general records, we observed four occurrences linked to other circumstances: three suicide attempts with repeated co-administration of medication and drugs of abuse (resulting in hospitalizations) and one accidental ingestion. We emphasize that even in these cases, everyone recovered completely.

With respect to annual incidents with psilocybin mushrooms, we observed four reports in 2022, four in 2021, three in 2019, one in 2015, and one in 2013. In contrast, the US recorded 4055 incidents involving psilocybin from 2013 to 2022, predominantly among teenagers and young adults aged 13-25 (Farah et al., 2024). This suggests potential underreporting of adverse events in Brazil, highlighting the need for ongoing regulatory discussions to ensure continuous monitoring.

Unfortunately, our ability to analyze some demographic characteristics is limited by the data collection system. SINAN notifications do not provide accurate information about education, pregnancy and exposure location. Therefore, we did not include it as part of the demographic characteristics of the participants. Another detail is that SINAN does not have records on income, risk behaviors, specific contexts of use, pre-existing pathological conditions, length of hospitalization, administration route, dose and frequency use of toxic agents. In the SINAN data, we observed a wide variation in the names of reported toxic agents, including multiple pieces of information in a single database cell, the use of street names for drugs, commercial names for medications, as well as typographical errors and other inaccurate nomenclatures.

Thus, we suggest the application of machine learning methods for greater accuracy and optimization of analysis time. This approach can facilitate ongoing surveillance worldwide. The general panorama of adverse events presented in our study must be interpreted based on the limitations presented. We hope that future studies will carry out more in-depth research.

## Conclusion

This study analyzed the profile of adverse events associated with drug abuse in Brazil from 2007 to 2022, with a focus on psilocybin mushrooms and unknown mushrooms based on SINAN data. Our findings indicate that, compared to other toxic agents, psilocybin mushrooms and unknown mushrooms account for a relatively small proportion of drug abuse incidents, with no fatalities documented during the study period. While our results suggest that adverse events related to psilocybin mushrooms are uncommon and have a low-risk profile, the possibility of underreporting cannot be ruled out. Further research using comprehensive monitoring methods is warranted. Given the increasing public interest in psilocybin mushrooms, we emphasize the importance of evidence-based regulatory discussions to prevent arbitrary arrests and ensure safe access to psilocybin for both clinical and ceremonial purposes.

## Supporting information

Appendix A. ANVISA's response.

Appendix B. Medications.

## Data Availability

All data produced in the present study are available upon reasonable request to the authors.

https://datasus.saude.gov.br/transferencia-de-arquivos

## Authority contribution statement

**Marcel Nogueira:** Writing - Original Draft, Writing – Review & Editing, Visualization, Validation, Supervision, Project administration, Methodology, Investigation, Software, Formal analysis, Data curation, Conceptualization. **Solimary García-Hernández:** Writing – Review & Editing, Visualization, Validation, Software, Formal analysis. **Gleicy Sotéro Roberto:** Writing – Review & Editing, Visualization, Validation, Software, Formal analysis. **Leonardo Marques Zanella:** Visualization, Software.

## Acknowledgment

Special thanks go to Daniel Cardoso Rodrigues for believing in our project. We would like to emphasize that he was not involved in the conduct of the research, data analysis, or writing of the manuscript.

