## Appendix A. ANVISA's response. for "Psilocybin mushrooms and public health in Brazil: a low-risk adverse event profile calls for evidence-based regulatory discussions"

DeepL

Subscribe to DeepL Pro to translate larger documents.
Visit www.DeepL.com/pro for more information.

#### Basic data of the demonstration

Type of Manifestation: Access to Information

Sphere: Federal

NUP: 25072.026294/2023-81

Recipient Body: ANVISA - National Health Surveillance Agency

Body of Interest:

Subject: Others in Health

Sub-subject:

Registration date: 10/05/2023

Situation: Completed

Deadline for reply: 12/06/2023

Incoming channel: Internet

Response mode: Through the system (with email alerts)

Registered By: Organ

Type of form: Access to information

Service:

Other Service:

#### Content of the Manifestation

Summary: Data on psilocybin, psilocin and psilocybe intoxication.

Content: I'd like to know more about psilocybin poisoning, psilocin and psilocybe mushroom sp.

Proposal for

improvement: Municipality

of the place of the event:

UF of the place of the

event:

Location:

There are no original annexes to the demonstration.

There are no additional annexes.

There are no supplementary texts.

There are no people involved in the demonstration.

#### Additional fields

### Integrated Platform for Ombudsman and Access to Information Details of the Manifestation

There are no additional fields.

### Integrated Platform for Ombudsman and Access to Information Details of the Manifestation

#### Response data

| Type of response | Date/Time | Response content | Decision |
|---|---|---|---|
| Conclusive Answer | 05/06/2023 12:19 | Dear Sir/Madam, In response to your request, and based on the information provided by the Controlled Products Management (GPCON), the technical area responsible for the matter in question, registered in the Anvisa Service System - SAT, Protocol No. 2023121893, we would like to inform you that Psylocibe cubensis is a species of mushroom whose composition includes the substances psilocybin and psilocin, both of which are banned in Brazil because they are classified on List F2 (List of psychotropic substances banned in Brazil) of Annex I of Ordinance SVS/MS No. 344/1998. There are no notifications of the substances psilocybin, psilocin and psilocibe in the databases used by the Pharmacovigilance Management - GFARM. We would like to inform you that the National Health Surveillance Agency has made available on its website, the Pharmacovigilance Notification Panel, which can be freely accessed via the following link https://www.gov.br/anvisa/pt-br/acessoainformacao/dadosabertos/informacoes-analytics/notifications-of-pharmacovigilance. In this panel, data on suspected adverse events related to the use of medicines can be consulted by citizens. The dashboard allows the public to access information on spontaneous notifications of suspected adverse drug and vaccine events received by Anvisa through VigiMed since December 2018, when the system was implemented. The data received in the VigiMed system can also be accessed through Brazilian Open Data Portal, available at https://dados.gov.br/dados/conjuntos-dados/notificacoes-em-farmacovigilancia. The notification data is presented in anonymously, without including personal data or data that could identify a person, in compliance with the provisions of the Access to Information Law (Law 12.527/2011) and the General Personal Data Protection Law (Law 13.709/2018). In compliance with the provisions of art. 11, § 4, of Law 12.527/11, we inform you that the applicant may file an appeal on Fala.BR - Integrated Platform for Ombudsman and Access to Information, within 10 (ten) days of receiving the decision, which will be evaluated by the General Management for Monitoring Products Subject to Sanitary Surveillance (GGMON). For further clarification, Anvisa also offers its Call Center, on 0800 642 9782 (working days, from 7:30 a.m. to 7:30 p.m.) and electronically, at Fale Conosco: (http://www.anvisa.gov.br/institucional/faleconosco/FaleConosco.asp) Sincerely, Agency National Health Surveillance | Access Granted |

### Integrated Platform for Ombudsman and Access to Information Details of the Manifestation

#### Reporting non-compliance

There are no reports of non-compliance.

#### Referral data

There are no records of referrals.

#### Extension data

| Original deadline | New Deadline | Responsible | Reason | Justification | Date/Time Action |
|---|---|---|---|---|---|
| 31/05/2023 23:59 | 12/06/2023 23:59 | Organ | Other reasons | Dear Sir or Madam,<br><br>We extend the response deadline because the area responsible technician needed a additional time, allowed by law 12.527/2011, the Access to Information (LAI). We apologize for the inconvenience and we thank understanding. Sincerely, Coordination of Management Transparency and Access to Information - CGTAI | 31/05/2023 16:44 |
