## Appendix B. Medications. for "Psilocybin mushrooms and public health in Brazil: a low-risk adverse event profile calls for evidence-based regulatory discussions"

**Appendix B. Anatomical Therapeutic Chemical (ATC) Classification**

**Medications – Overall**

| **N** | **14,794** | | **p-value** |
|---|---|---|---|
| **ATC 2nd and 3rd levels** | **N** | **(%)** | |
| A02 DRUGS FOR ACID RELATED DISORDERS | 150 | 1.0 | <0.001 |
| A03 DRUGS FOR FUNCTIONAL GASTROINTESTINAL DISORDERS | 74 | 0.5 | |
| A04 ANTIEMETICS AND ANTINAUSEANTS | 74 | 0.5 | |
| A06 DRUGS FOR CONSTIPATION | 13 | 0.1 | |
| A07 ANTIDIARRHEALS. INTESTINAL ANTIINFLAMMATORY/ANTIINFECTIVE AGENTS | 1 | 0.0 | |
| A08 ANTIOBESITY PREPARATIONS. EXCL. DIET PRODUCTS | 51 | 0.3 | |
| A10 DRUGS USED IN DIABETES | 94 | 0.6 | |
| A11 VITAMINS | 62 | 0.4 | |
| A12 MINERAL SUPPLEMENTS | 11 | 0.1 | |
| A14 ANABOLIC AGENTS FOR SYSTEMIC USE | 7 | 0.0 | |
| B01A ANTITHROMBOTIC AGENTS | 13 | 0.1 | |
| B02 ANTIHEMORRHAGICS | 2 | 0.0 | |
| B03 ANTIANEMIC PREPARATIONS | 18 | 0.1 | |
| C01 CARDIAC THERAPY | 15 | 0.1 | |
| C01C CARDIAC STIMULANTS EXCL. CARDIAC GLYCOSIDES | 7 | 0.0 | |
| C02 ANTIHYPERTENSIVES | 21 | 0.1 | |
| C03 DIURETICS | 77 | 0.5 | |
| C05 VASOPROTECTIVES | 2 | 0.0 | |
| C07A BETA BLOCKING AGENTS | 83 | 0.6 | |
| C08 CALCIUM CHANNEL BLOCKERS | 22 | 0.1 | |
| C09A ACE INHIBITORS. PLAIN | 145 | 1.0 | |

| | | |
|---|---|---|
| C09C ANGIOTENSIN II RECEPTOR BLOCKERS (ARBs). PLAIN | 87 | 0.6 |
| C10 LIPID MODIFYING AGENTS | 30 | 0.2 |
| D02 EMOLLIENTS AND PROTECTIVES | 2 | 0.0 |
| D08 ANTISEPTICS AND DISINFECTANTS | 7 | 0.0 |
| D10 ANTI-ACNE PREPARATIONS | 1 | 0.0 |
| G02A UTEROTONICS | 1 | 0.0 |
| G03A HORMONAL CONTRACEPTIVES FOR SYSTEMIC USE | 35 | 0.2 |
| G03H ANTIANDROGENS | 1 | 0.0 |
| G03B ANDROGENS | 46 | 0.3 |
| G04B UROLOGICALS | 7 | 0.0 |
| G04BE Drugs used in erectile dysfunction | 33 | 0.2 |
| H01 PITUITARY AND HYPOTHALAMIC HORMONES AND ANALOGUES | 2 | 0.0 |
| H02 CORTICOSTEROIDS FOR SYSTEMIC USE | 36 | 0.2 |
| H03 THYROID THERAPY | 14 | 0.1 |
| J05 ANTIVIRALS FOR SYSTEMIC USE | 18 | 0.1 |
| J01A TETRACYCLINES | 10 | 0.1 |
| J02 ANTIMYCOTICS FOR SYSTEMIC USE | 10 | 0.1 |
| J04A DRUGS FOR TREATMENT OF TUBERCULOSIS | 4 | 0.0 |
| J01C BETA-LACTAM ANTIBACTERIALS. PENICILLINS | 124 | 0.8 |
| J01D OTHER BETA-LACTAM ANTIBACTERIALS | 58 | 0.4 |
| J01E SULFONAMIDES AND TRIMETHOPRIM | 14 | 0.1 |
| J01F MACROLIDES. LINCOSAMIDES AND STREPTOGRAMINS | 19 | 0.1 |
| J01M QUINOLONE ANTIBACTERIALS | 43 | 0.3 |
| J01X OTHER ANTIBACTERIALS | 1 | 0.0 |
| J04B DRUGS FOR TREATMENT OF LEPRA | 2 | 0.0 |
| L02 ENDOCRINE THERAPY | 2 | 0.0 |
| L04 IMMUNOSUPPRESSANTS | 2 | 0.0 |
| M01A ANTIINFLAMMATORY AND ANTIRHEUMATIC PRODUCTS. NON-STEROIDS | 451 | 3.0 |

| | | |
|---|---|---|
| N01 ANESTHETICS | 3 | 0.0 |
| M03 MUSCLE RELAXANTS | 224 | 1.5 |
| M04A ANTIGOUT PREPARATIONS | 2 | 0.0 |
| N02A OPIOIDS | 189 | 1.3 |
| N02B OTHER ANALGESICS AND ANTIPYRETICS | 880 | 5.9 |
| N02C ANTIMIGRAINE PREPARATIONS | 8 | 0.1 |
| N03A ANTIEPILEPTICS | 4769 | 32.2 |
| N04 ANTI-PARKINSON DRUGS | 64 | 0.4 |
| N05A ANTIPSYCHOTICS | 1251 | 8.5 |
| N05B ANXIOLYTICS | 1834 | 12.4 |
| N05C HYPNOTICS AND SEDATIVES | 324 | 2.2 |
| N06A ANTIDEPRESSANTS | 2271 | 15.4 |
| N06B PSYCHOSTIMULANTS. AGENTS USED FOR ADHD AND NOOTROPICS | 322 | 2.2 |
| N07B DRUGS USED IN ADDICTIVE DISORDERS | 41 | 0.3 |
| N07C ANTIVERTIGO PREPARATIONS | 21 | 0.1 |
| P01 ANTIPROTOZOALS | 26 | 0.2 |
| P01B ANTIMALARIALS | 2 | 0.0 |
| P02C ANTINEMATODAL AGENTS | 13 | 0.1 |
| P03 ECTOPARASITICIDES. INCL. SCABICIDES. INSECTICIDES AND REPELLENTS | 28 | 0.2 |
| R01 NASAL PREPARATIONS | 29 | 0.2 |
| R03 DRUGS FOR OBSTRUCTIVE AIRWAY DISEASES | 68 | 0.5 |
| R05 COUGH AND COLD PREPARATIONS | 4 | 0.0 |
| R06A ANTIHISTAMINES FOR SYSTEMIC USE | 392 | 2.6 |
| S01 OPHTHALMOLOGICALS | 2 | 0.0 |
| S02 OTOLOGICALS | 3 | 0.0 |
| V03 ALL OTHER THERAPEUTIC PRODUCTS | 22 | 0.1 |

### Medications – Non-hospitalization

| N | 10,988 | | p-value |
|---|---|---|---|
| **ATC 2nd and 3rd levels** | **N** | **(%)** | |
| A02 DRUGS FOR ACID RELATED DISORDERS | 104 | 0.9 | <0.001 |
| A03 DRUGS FOR FUNCTIONAL GASTROINTESTINAL DISORDERS | 54 | 0.5 | |
| A04 ANTIEMETICS AND ANTINAUSEANTS | 61 | 0.5 | |
| A06 DRUGS FOR CONSTIPATION | 12 | 0.1 | |
| A07 ANTIDIARRHEALS. INTESTINAL ANTIINFLAMMATORY/ANTIINFECTIVE AGENTS | 1 | 0.0 | |
| A08 ANTIOBESITY PREPARATIONS. EXCL. DIET PRODUCTS | 41 | 0.4 | |
| A10 DRUGS USED IN DIABETES | 59 | 0.5 | |
| A11 VITAMINS | 44 | 0.4 | |
| A12 MINERAL SUPPLEMENTS | 6 | 0.0 | |
| A14 ANABOLIC AGENTS FOR SYSTEMIC USE | 4 | 0.0 | |
| B01A ANTITHROMBOTIC AGENTS | 5 | 0.0 | |
| B02 ANTIHEMORRHAGICS | 1 | 0.0 | |
| B03 ANTIANEMIC PREPARATIONS | 15 | 0.1 | |
| C01 CARDIAC THERAPY | 8 | 0.0 | |
| C01C CARDIAC STIMULANTS EXCL. CARDIAC GLYCOSIDES | 6 | 0.0 | |
| C02 ANTIHYPERTENSIVES | 14 | 0.1 | |
| C03 DIURETICS | 58 | 0.5 | |
| C05 VASOPROTECTIVES | 2 | 0.0 | |
| C07A BETA BLOCKING AGENTS | 52 | 0.5 | |
| C08 CALCIUM CHANNEL BLOCKERS | 13 | 0.1 | |
| C09A ACE INHIBITORS. PLAIN | 97 | 0.9 | |
| C09C ANGIOTENSIN II RECEPTOR BLOCKERS (ARBs). PLAIN | 65 | 0.6 | |
| C10 LIPID MODIFYING AGENTS | 20 | 0.2 | |
| D02 EMOLLIENTS AND PROTECTIVES | 2 | 0.0 | |

| | | |
|---|---|---|
| D08 ANTISEPTICS AND DISINFECTANTS | 3 | 0.0 |
| D10 ANTI-ACNE PREPARATIONS | 1 | 0.0 |
| H02 CORTICOSTEROIDS FOR SYSTEMIC USE | 35 | 0.3 |
| G02A UTEROTONICS | 1 | 0.0 |
| G03A HORMONAL CONTRACEPTIVES FOR SYSTEMIC USE | 35 | 0.3 |
| G03H ANTIANDROGENS | 1 | 0.0 |
| G03B ANDROGENS | 29 | 0.3 |
| G04B UROLOGICALS | 5 | 0.0 |
| G04BE Drugs used in erectile dysfunction | 15 | 0.1 |
| H03 THYROID THERAPY | 11 | 0.1 |
| J05 ANTIVIRALS FOR SYSTEMIC USE | 15 | 0.1 |
| J01A TETRACYCLINES | 7 | 0.0 |
| J02 ANTIMYCOTICS FOR SYSTEMIC USE | 8 | 0.0 |
| J04A DRUGS FOR TREATMENT OF TUBERCULOSIS | 4 | 0.0 |
| J01C BETA-LACTAM ANTIBACTERIALS. PENICILLINS | 95 | 0.8 |
| J01D OTHER BETA-LACTAM ANTIBACTERIALS | 44 | 0.4 |
| J01E SULFONAMIDES AND TRIMETHOPRIM | 14 | 0.1 |
| J01F MACROLIDES, LINCOSAMIDES AND STREPTOGRAMINS | 17 | 0.1 |
| J01M QUINOLONE ANTIBACTERIALS | 35 | 0.3 |
| J01X OTHER ANTIBACTERIALS | 1 | 0.0 |
| J04B DRUGS FOR TREATMENT OF LEPRA | 2 | 0.0 |
| L02 ENDOCRINE THERAPY | 1 | 0.0 |
| M01A ANTIINFLAMMATORY AND ANTIRHEUMATIC PRODUCTS. NON-STEROIDS | 350 | 3.1 |
| N01 ANESTHETICS | 1 | 0.0 |
| M03 MUSCLE RELAXANTS | 172 | 1.6 |
| M04A ANTIGOUT PREPARATIONS | 2 | 0.0 |
| N02A OPIOIDS | 110 | 1.0 |
| N02B OTHER ANALGESICS AND ANTIPYRETICS | 675 | 6.1 |

| | | |
|---|---|---|
| N02C ANTIMIGRAINE PREPARATIONS | 7 | 0.0 |
| N03A ANTIEPILEPTICS | 3341 | 30.4 |
| N04 ANTI-PARKINSON DRUGS | 40 | 0.4 |
| N05A ANTIPSYCHOTICS | 836 | 7.7 |
| N05B ANXIOLYTICS | 1805 | 16.4 |
| N05C HYPNOTICS AND SEDATIVES | 198 | 1.8 |
| N06A ANTIDEPRESSANTS | 1587 | 14.4 |
| N06B PSYCHOSTIMULANTS. AGENTS USED FOR ADHD AND NOOTROPICS | 256 | 2.3 |
| N07B DRUGS USED IN ADDICTIVE DISORDERS | 21 | 0.2 |
| N07C ANTIVERTIGO PREPARATIONS | 18 | 0.2 |
| P01 ANTIPROTOZOALS | 21 | 0.2 |
| P01B ANTIMALARIALS | 1 | 0.0 |
| P02C ANTINEMATODAL AGENTS | 10 | 0.0 |
| P03 ECTOPARASITICIDES. INCL. SCABICIDES. INSECTICIDES AND REPELLENTS | 17 | 0.1 |
| R01 NASAL PREPARATIONS | 22 | 0.2 |
| R03 DRUGS FOR OBSTRUCTIVE AIRWAY DISEASES | 61 | 0.5 |
| R05 COUGH AND COLD PREPARATIONS | 3 | 0.0 |
| R06A ANTIHISTAMINES FOR SYSTEMIC USE | 289 | 2.6 |
| S01 OPHTHALMOLOGICALS | 2 | 0.0 |
| S02 OTOLOGICALS | 3 | 0.0 |
| V03 ALL OTHER THERAPEUTIC PRODUCTS | 17 | 0.1 |

**Medications – Hospitalization**

| N | 3654 | | p-value |
|---|---|---|---|
| **ATC 2nd and 3rd levels** | **N** | **(%)** | |
| A02 DRUGS FOR ACID RELATED DISORDERS | 46 | 1.3 | <0.001 |
| A03 DRUGS FOR FUNCTIONAL GASTROINTESTINAL DISORDERS | 20 | 0.5 | |
| A04 ANTIEMETICS AND ANTINAUSEANTS | 13 | 0.4 | |
| A06 DRUGS FOR CONSTIPATION | 1 | 0.0 | |
| A08 ANTIOBESITY PREPARATIONS. EXCL. DIET PRODUCTS | 10 | 0.3 | |
| A10 DRUGS USED IN DIABETES | 34 | 0.9 | |
| A11 VITAMINS | 18 | 0.5 | |
| A12 MINERAL SUPPLEMENTS | 5 | 0.1 | |
| A14 ANABOLIC AGENTS FOR SYSTEMIC USE | 3 | 0.1 | |
| B01A ANTITHROMBOTIC AGENTS | 8 | 0.2 | |
| B02 ANTIHEMORRHAGICS | 1 | 0,0 | |
| B03 ANTIANEMIC PREPARATIONS | 3 | 0.1 | |
| C01 CARDIAC THERAPY | 7 | 0.2 | |
| C01C CARDIAC STIMULANTS EXCL. CARDIAC GLYCOSIDES | 1 | 0.0 | |
| C02 ANTIHYPERTENSIVES | 7 | 0.2 | |
| C03 DIURETICS | 19 | 0.5 | |
| C07A BETA BLOCKING AGENTS | 30 | 0.8 | |
| C08 CALCIUM CHANNEL BLOCKERS | 9 | 0.2 | |
| C09A ACE INHIBITORS. PLAIN | 45 | 1.2 | |
| C09C ANGIOTENSIN II RECEPTOR BLOCKERS (ARBs). PLAIN | 22 | 0.6 | |
| C10 LIPID MODIFYING AGENTS | 10 | 0.3 | |
| D08 ANTISEPTICS AND DISINFECTANTS | 4 | 0.1 | |
| H02 CORTICOSTEROIDS FOR SYSTEMIC USE | 1 | 0.0 | |
| G03B ANDROGENS | 17 | 0.5 | |

| | | |
|---|---|---|
| G04B UROLOGICALS | 2 | 0.1 |
| G04BE Drugs used in erectile dysfunction | 12 | 0.3 |
| H01 PITUITARY AND HYPOTHALAMIC HORMONES AND ANALOGUES | 1 | 0.0 |
| H03 THYROID THERAPY | 3 | 0.1 |
| J05 ANTIVIRALS FOR SYSTEMIC USE | 2 | 0.1 |
| J01A TETRACYCLINES | 3 | 0.1 |
| J02 ANTIMYCOTICS FOR SYSTEMIC USE | 2 | 0.1 |
| J01C BETA-LACTAM ANTIBACTERIALS. PENICILLINS | 29 | 0.8 |
| J01D OTHER BETA-LACTAM ANTIBACTERIALS | 14 | 0.4 |
| J01F MACROLIDES. LINCOSAMIDES AND STREPTOGRAMINS | 2 | 0.1 |
| J01M QUINOLONE ANTIBACTERIALS | 8 | 0.2 |
| L02 ENDOCRINE THERAPY | 1 | 0.0 |
| L04 IMMUNOSUPPRESSANTS | 2 | 0.1 |
| M01A ANTIINFLAMMATORY AND ANTIRHEUMATIC PRODUCTS. NON-STEROIDS | 96 | 2.6 |
| N01 ANESTHETICS | 1 | 0.0 |
| M03 MUSCLE RELAXANTS | 51 | 1.4 |
| M04A ANTIGOUT PREPARATIONS | 0 | 0.0 |
| N02A OPIOIDS | 68 | 1.9 |
| N02B OTHER ANALGESICS AND ANTIPYRETICS | 202 | 5.5 |
| N02C ANTIMIGRAINE PREPARATIONS | 1 | 0.0 |
| N03A ANTIEPILEPTICS | 1386 | 37.9 |
| N04 ANTI-PARKINSON DRUGS | 22 | 0.6 |
| N05A ANTIPSYCHOTICS | 401 | 11.0 |
| N05B ANXIOLYTICS | 2 | 0.1 |
| N05C HYPNOTICS AND SEDATIVES | 122 | 3.3 |
| N06A ANTIDEPRESSANTS | 662 | 18.1 |
| N06B PSYCHOSTIMULANTS. AGENTS USED FOR ADHD AND NOOTROPICS | 66 | 1.8 |
| N07B DRUGS USED IN ADDICTIVE DISORDERS | 20 | 0.5 |

| | | |
|---|---|---|
| N07C ANTIVERTIGO PREPARATIONS | 3 | 0.1 |
| P01 ANTIPROTOZOALS | 5 | 0.1 |
| P01B ANTIMALARIALS | 1 | 0.0 |
| P02C ANTINEMATODAL AGENTS | 2 | 0.1 |
| P03 ECTOPARASITICIDES. INCL. SCABICIDES. INSECTICIDES AND REPELLENTS | 11 | 0.3 |
| R01 NASAL PREPARATIONS | 7 | 0.2 |
| R03 DRUGS FOR OBSTRUCTIVE AIRWAY DISEASES | 7 | 0.2 |
| R06A ANTIHISTAMINES FOR SYSTEMIC USE | 99 | 2.7 |
| V03 ALL OTHER THERAPEUTIC PRODUCTS | 4 | 4.0 |

**Medications – Death**

| N | 152 | | p-value |
|---|---|---|---|
| **ATC 2nd and 3rd levels** | **N** | **(%)** | |
| A10 DRUGS USED IN DIABETES | 1 | 0.7 | |
| C07A BETA BLOCKING AGENTS | 1 | 0.7 | |
| C09A ACE INHIBITORS. PLAIN | 3 | 2.0 | |
| G04BE Drugs used in erectile dysfunction | 6 | 3.9 | |
| H01 PITUITARY AND HYPOTHALAMIC HORMONES AND ANALOGUES | 1 | 0.7 | |
| J05 ANTIVIRALS FOR SYSTEMIC USE | 1 | 0.7 | |
| M01A ANTIINFLAMMATORY AND ANTIRHEUMATIC PRODUCTS. NON-STEROIDS | 5 | 3.3 | |
| N01 ANESTHETICS | 1 | 0.7 | |
| M03 MUSCLE RELAXANTS | 1 | 0.7 | |
| N02A OPIOIDS | 11 | 7.2 | <0.001 |
| N02B OTHER ANALGESICS AND ANTIPYRETICS | 3 | 2.0 | |
| N03A ANTIEPILEPTICS | 42 | 27.6 | |
| N04 ANTI-PARKINSON DRUGS | 2 | 1.3 | |
| N05A ANTIPSYCHOTICS | 14 | 9.2 | |
| N05B ANXIOLYTICS | 27 | 17.8 | |
| N05C HYPNOTICS AND SEDATIVES | 4 | 2.6 | |
| N06A ANTIDEPRESSANTS | 22 | 14.5 | |
| P02C ANTINEMATODAL AGENTS | 1 | 0.7 | |
| R05 COUGH AND COLD PREPARATIONS | 1 | 0.7 | |
| R06A ANTIHISTAMINES FOR SYSTEMIC USE | 4 | 2.6 | |
| V03 ALL OTHER THERAPEUTIC PRODUCTS | 1 | 0.7 | |

| ATC Classification | Identified medications |
|---|---|
| A02 DRUGS FOR ACID RELATED DISORDERS | cimetidine, famotidine, dexlansoprazole, ranitidine, omeprazole, pantoprazole, vonoprazan, sodium bicarbonate, aluminum hydroxide |
| A03 DRUGS FOR FUNCTIONAL GASTROINTESTINAL DISORDERS | bromopride, domperidone, isometheptene, metoclopramide, atropine, papaverine, hyoscyamine, piperidolate |
| A04 ANTIEMETICS AND ANTINAUSEANTS | scopolamine, ondansetron |
| A06 DRUGS FOR CONSTIPATION | bisacodyl, mineral oil |
| A07 ANTIDIARRHEALS, INTESTINAL ANTIINFLAMMATORY/ANTIINFECTIVE AGENTS | loperamide |
| A08 ANTIOBESITY PREPARATIONS, EXCL. DIET PRODUCTS | amfepramone, sibutramine |
| A10 DRUGS USED IN DIABETES | alogliptin, metformin, gliclazide, insulin, glibenclamide, dapagliflozin |
| A11 VITAMINS | ascorbic acid, B vitamins, fat-soluble vitamins |
| A12 MINERAL SUPPLEMENTS | calcium, potassium chloride, sodium chloride, magnesium, zinc |
| A14 ANABOLIC AGENTS FOR SYSTEMIC USE | oxandrolone, trenbolone, stanozolol, nandrolone, oxymetholone |
| B01A ANTITHROMBOTIC AGENTS | cilostazol, rivaroxaban, warfarin |
| B02 ANTIHEMORRHAGICS | tranexamic acid |
| B03 ANTIANEMIC PREPARATIONS | iron, folic acid, vitamin b12, ferripolymaltose, erythropoietin |
| C01 CARDIAC THERAPY | amiodarone, propafenone, digoxin, isosorbide, propatyl nitrate |
| C01C CARDIAC STIMULANTS EXCL. CARDIAC GLYCOSIDES | ephedrine, adrenaline, mephentermine |
| C02 ANTIHYPERTENSIVES | hydralazine, clonidine, methyldopa |
| C03 DIURETICS | hydrochlorothiazide, furosemide, chlorthalidone, spironolactone |
| C05 VASOPROTECTIVES | troxerutin, coumarin |
| C07A BETA BLOCKING AGENTS | atenolol, metoprolol, carvedilol, propranolol |
| C08 CALCIUM CHANNEL BLOCKERS | nimodipine, nifedipine, amlodipine |

| | |
|---|---|
| C09A ACE INHIBITORS, PLAIN | captopril, enalapril maleate, ramipril |
| C09C ANGIOTENSIN II RECEPTOR BLOCKERS (ARBS), PLAIN | losartan, valsartan |
| C10 LIPID MODIFYING AGENTS | atorvastatin, simvastatin, rosuvastatin |
| D02 EMOLLIENTS AND PROTECTIVES | salicylic acid |
| D08 ANTISEPTICS AND DISINFECTANTS | benzalkonium chloride, nitrofurazone |
| D10 ANTI-ACNE PREPARATIONS | isotretinoin |
| H02 CORTICOSTEROIDS FOR SYSTEMIC USE | betamethasone, prednisone, dexamethasone, prednisolone |
| G02A UTEROTONICS | methylergometrine |
| G03A HORMONAL CONTRACEPTIVES FOR SYSTEMIC USE | levonorgestrel, ethinylestradiol, drospirenone, gestodene, norethisterone |
| G03H ANTIANDROGENS | cyproterone |
| G03B ANDROGENS | testosterone, fluoxymesterone |
| G04B UROLOGICALS | phenazopyridine, oxybutynin, methenamine, acriflavine |
| G04BE DRUGS USED IN ERECTILE DYSFUNCTION | sildenafil, tadalafil |
| H01 PITUITARY AND HYPOTHALAMIC HORMONES AND ANALOGUES | somatropin |
| H03 THYROID THERAPY | levothyroxine, tapazole |
| J05 ANTIVIRALS FOR SYSTEMIC USE | zidovudine, lamivudine, efavirenz, acyclovir |
| J01A TETRACYCLINES | tetracycline, doxycycline |
| J02 ANTIMYCOTICS FOR SYSTEMIC USE | fluconazole, ketoconazole, chloramphenicol |
| J04A DRUGS FOR TREATMENT OF TUBERCULOSIS | isoniazid, rifampicin |
| J01C BETA-LACTAM ANTIBACTERIALS, PENICILLINS | amoxicillin, clavulanate, ampicillin, oxacillin, benzylpenicillin |
| J01D OTHER BETA-LACTAM ANTIBACTERIALS | cefuroxime axetil, cephalexin, cefadroxil |
| J01E SULFONAMIDES AND TRIMETHOPRIM | sulfamethoxazole, trimethoprim, sulfadiazine |

| | |
|---|---|
| J01F MACROLIDES, LINCOSAMIDES AND STREPTOGRAMINS | azithromycin, erythromycin |
| J01M QUINOLONE ANTIBACTERIALS | ciprofloxacin, enrofloxacin, levofloxacin, norfloxacin, nalidixic acid |
| J01X OTHER ANTIBACTERIALS | nitrofurantoin |
| J04B DRUGS FOR TREATMENT OF LEPRA | dapsone |
| L02 ENDOCRINE THERAPY | tamoxifen |
| L04 IMMUNOSUPPRESSANTS | mycophenolate, tacrolimus |
| M01A ANTIINFLAMMATORY AND ANTIRHEUMATIC PRODUCTS, NON-STEROIDS | ibuprofen, naproxen, meloxicam, piroxicam, nimesulide, diclofenac, ketoprofen, ketorolac, mefenamic acid, benzydamine, celecoxib, etoricoxib, etodolac, tenoxicam |
| N01 ANESTHETICS | lidocaine, procaine, etomidate, tetracaine |
| M03 MUSCLE RELAXANTS | baclofen, cyclobenzaprine, carisoprodol, orphenadrine, adifenine |
| M04A ANTIGOUT PREPARATIONS | colchicine, thiocolchicoside |
| N02A OPIOIDS | oxycodone, morphine, tramadol, codeine, pethidine |
| N02B OTHER ANALGESICS AND ANTIPYRETICS | metamizole (dipyrone), paracetamol, AAS |
| N02C ANTIMIGRAINE PREPARATIONS | dihydroergotamine, ergotamine, sumatriptan, naratriptan |
| N03A ANTIEPILEPTICS | carbamazepine, cannabidiol, topiramate, gabapentin, pregabalin, levetiracetam, lamotrigine, phenytoin, phenobarbital, clonazepam, oxcarbazepine, primidone, valproic acid, sodium valproate |
| N04 ANTI-PARKINSON DRUGS | biperiden, levodopa, trihexyphenidyl, benserazide, amantadine |
| N05A ANTIPSYCHOTICS | chlorpromazine, thioridazine, lurasidone, ziprasidone, trifluoperazine, periciazine, pimozide, risperidone, haloperidol, quetiapine, sulpiride, lithium, aripiprazole, clozapine, olanzapine, levomepromazine |
| N05B ANXIOLYTICS | cloxazolam, alprazolam, diazepam, bromazepam, clobazam, chlordiazepoxide, lorazepam |
| N05C HYPNOTICS AND SEDATIVES | flunitrazepam, zolpidem, estazolam, zopiclone, eszopiclone, flurazepam, midazolam, melatonin, nitrazepam |
| N06A ANTIDEPRESSANTS | amitriptyline, agomelatine, maprotiline, clomipramine, vortioxetine, imipramine, fluoxetine, fluvoxamine, escitalopram, citalopram, paroxetine, duloxetine, venlafaxine, mirtazapine, trazodone, bupropion, buspirone |

| | |
|---|---|
| N06B PSYCHOSTIMULANTS, AGENTS USED FOR ADHD AND NOOTROPICS | caffeine, guarana, methylphenidate, lisdexamfetamine |
| N07B DRUGS USED IN ADDICTIVE DISORDERS | disulfiram, methadone, naltrexone |
| N07C ANTIVERTIGO PREPARATIONS | betahistine, cinnarizine, flunarizine |
| P01 ANTIPROTOZOALS | metronidazole |
| P01B ANTIMALARIALS | chloroquine |
| P02C ANTINEMATODAL AGENTS | ivermectin, nitazoxanide, mebendazole, piperazine, secnidazole, thiabendazole, levamisole |
| P03 ECTOPARASITICIDES, INCL. SCABICIDES, INSECTICIDES AND REPELLENTS | dimethicone, permethrin, deltamethrin |
| R01 NASAL PREPARATIONS | phenylephrine, naphazoline, pseudoephedrine |
| R03 DRUGS FOR OBSTRUCTIVE AIRWAY DISEASES | fenoterol, ipratropium, aminophylline, budesonide, clenbuterol, formoterol, salbutamol, theophylline, montelukast, terbutaline |
| R05 COUGH AND COLD PREPARATIONS | acebrophylline, acetylcysteine, ambroxol, bromhexine, levodropropizine, clobutinol, guaifenesin, oxymetazoline, potassium iodide |
| R06A ANTIHISTAMINES FOR SYSTEMIC USE | diphenhydramine, fexofenadine, brompheniramine, meclizine, carbinoxamine, dimenhydrinate, doxylamine, promethazine, hydroxyzine, loratadine, buclizine, cyproheptadine, dexchlorpheniramine, chlorpheniramine, clemastine |
| S01 OPHTHALMOLOGICALS | brimonidine, hypromellose |
| S02 OTOLOGICALS | boric acid |
| V03 ALL OTHER THERAPEUTIC PRODUCTS | methionine, racemethionine, silymarin, choline, betaine, phenylethylamine, l-arginine, goji berry, bacterial immunostimulant, potassium permanganate |
